# The role of EEG in predicting post-stroke seizures and an updated prognostic model (SeLECT-EEG)

**DOI:** 10.1101/2024.11.28.24318126

**Authors:** Kai Michael Schubert, Vijaya Dasari, Ana Lúcia Oliveira, Chiara Tatillo, Gilles Naeije, Adam Strzelczyk, Nicolas Gaspard, Marian Galovic, Vineet Punia, Carla Bentes, the SeLECT Collaborators

## Abstract

**Importance:** Seizures significantly impact outcomes after stroke, underscoring the need for accurate predictors of post-stroke epilepsy.

**Objective:** To evaluate whether electrographic biomarkers detected early after acute ischemic stroke enhance the prediction of post-stroke epilepsy.

**Design:** Multicenter cohort study with data collected from 2002 to 2022 and final data analysis completed in July 2024.

**Setting:** Eleven international cohorts from tertiary referral centers, six with available EEG data.

**Participants:** 1,105 stroke survivors with neuroimaging-confirmed ischemic stroke (mean age 71, 54% male) who underwent EEG within the first 7 days post-stroke.

**Exposure:** Presence of electrographic biomarkers detected through EEG.

**Main Outcome and Measures:** Occurrence of post-stroke epilepsy. The impact of electrographic biomarkers on the risk of post-stroke epilepsy was assessed using Cox proportional hazards regression, adjusted through inverse probability weighting.

**Results:** Among 1,105 participants, 119 (11%) developed post-stroke seizures. Epileptiform activity (lateralized periodic discharges, interictal epileptiform discharges, and electrographic seizures; (odds ratio [OR] 2.0, 95% confidence interval [CI]: 1.3-3.0, p=0.001)) and regional slowing (OR 1.9, 95% CI: 1.2-2.9, p=0.004) were independently associated with developing post-stroke epilepsy. The novel SeLECT-EEG prognostic model, specifically developed for stroke survivors without acute symptomatic seizures (ASyS),, outperformed the previous gold-standard model (SeLECT_2.0_; 0.71 [95% CI: 0.65-0.76]) with a concordance statistic of 0.75 (95% CI: 0.71-0.80; p < 0.001).

**Conclusions and Relevance:** Electrographic findings significantly enhance the prediction of post-stroke epilepsy beyond previously known clinical risk factors and may serve as prognostic biomarkers. The integration of these biomarkers into the SeLECT-EEG model in patients without acute symptomatic seizures provides a more accurate prognostic tool for early post-stroke epilepsy prediction.

**Key points:** *Question:* Can early detection of electrographic biomarkers after acute ischemic stroke improve the prediction of post-stroke epilepsy?

*Findings:* Among 1,105 stroke survivors who received early EEG (≤ 7 days after stroke), post-stroke seizures occurred in 119 (11%). Stroke survivors with epileptiform activity had a 42% risk (95% CI 30%-49%) of developing post-stroke epilepsy 5 years after stroke, compared to a 13% risk (95% CI 9%-16%) in those without. Additionally, the 5-year risk of post-stroke epilepsy was twice as high in those with regional slowing (24%, 95% CI 18%-29%) compared to those without it (11%, 95% CI 5%-15%). Beyond known clinical risk factors, epileptiform activity and regional slowing were independently associated with developing post-stroke epilepsy. We integrated these findings into a novel prognostic model (SeLECT-EEG; concordance statistic 0.75 [95% CI: 0.71-0.80]), which outperformed the previous gold-standard model (SeLECT2.0; concordance statistic 0.71 [95% CI: 0.65-0.76]; p < 0.001).

*Meaning:* Early electrographic biomarkers improve the prediction of post-stroke epilepsy and may inform counseling and management strategies for stroke survivors at risk of seizures.

## Introduction

Stroke is the leading cause of epilepsy in older adults, accounting for over half of new-onset epilepsy cases in individuals aged 65 and above.^1^ Post-stroke seizures increase the risk of mortality, poor functional outcomes, disability, and dementia.^2-5^ Identifying accurate early predictors of post-stroke epilepsy is crucial for developing individualized management strategies for stroke survivors.

Although ASyS significantly increase the risk of post-stroke epilepsy,^6^ they occur in only a minority of patients, making it crucial to develop better predictive models and explore additional modalities for the majority of stroke patients without ASyS who are still at risk of developing post-stroke epilepsy.

Despite epilepsy being an electrophysiological disorder, the role of early EEG findings in predicting post-stroke epilepsy remains underexplored. Known predictors primarily include clinical and neuroimaging parameters, such as acute symptomatic (≤7 days post-stroke) clinical seizures, seizure type and timing,^4, 5^ stroke severity, large-artery atherosclerosis, cortical involvement, middle cerebral artery (MCA) territory involvement, and hemorrhagic conversion.^6, 7^ These factors are incorporated into clinical scores like SeLECT, SeLECT 2.0, and SeLECT-ASyS prognostic models (Schubert et al. 2024, under review).^4, 8^

Use of EEG in managing patients with acute brain injuries like stroke is already prevalent in the U.S. and is gaining popularity in Europe.^9^ Up to 12% of acute ischemic stroke survivors present with electrographic seizures (ESz), and 25% show rhythmic or periodic patterns (RPPs) on EEG during the acute phase.^10-13^ Risk factors for ESz and RPPs are similar to those for post-stroke epilepsy, suggesting they may herald post-stroke epilepsy.^14^ EEG is also sensitive to ischemic cortical injury and has been found to predict post-stroke epilepsy following other brain insults like traumatic brain injury (TBI).^15-17^ Continuous EEG (cEEG) monitoring has shown that highly epileptogenic RPPs are associated with the development of post-stroke epilepsy and can likely complement clinical risk factors.^18, 19^

This study aims to clarify whether electrographic findings on short, commonly used, and widely available scalp EEG, as well as cEEG during the acute phase of ischemic stroke, are associated with a higher risk of post-stroke epilepsy. It also seeks to determine if these EEG findings provide complementary prognostic information to clinical risk factors and help improve currently available prognostic models, particularly for patients without ASyS.

## Methods

### Participants

The derivation cohort included 980 participants from five international cohorts. The validation cohort included 125 participants. All participants were adults with acute ischemic stroke who received EEG within the first 7 days after stroke. For inverse probability weighting, we additionally analysed data from a multicenter registry of seizures after ischemic stroke established as part of the SeLECT study,^8^ (eight international cohorts comprising 3,711 participants). Individuals with transient ischemic attacks, a previous history of seizures or epilepsy, primary hemorrhagic stroke, re-infarction during follow-up, or potentially epileptogenic comorbidities (such as intracranial tumors, cerebral venous thrombosis, severe traumatic brain injury, or prior brain surgery) were excluded from the study. Detailed descriptions of the individual cohorts are provided in the Online Supplement. Informed consent was obtained either in written or verbal form. Four cohorts utilized written consent, while two cohorts employed both written and verbal consent. In five cohorts, consent requirements were waived by the regulatory authorities, as detailed in the online supplement.

### Definitions

Seizures were classified as acute symptomatic seizures (ASyS) if they occurred within 7 days of the stroke and remote symptomatic seizures (RSyS) if they were unprovoked and occurred more than 7 days after the stroke, in accordance with ILAE guidelines (Beghi 2011). The occurrence of RSyS was categorized as post-stroke epilepsy due to its high risk of seizure recurrence, exceeding the 60% risk required for the ILAE epilepsy definition.^20^ Status epilepticus was classified according to the revised ILAE definition,^21^ and electrographic status epilepticus was defined based on the Salzburg criteria.^22^

Short EEG recordings were defined as those lasting between 20 to 60 minutes, while cEEG recordings lasted at least 12 hours. Each cEEG was preceded by a 20-minute EEG screening. Epileptiform activity on EEG included electrographic seizures (ESz) and electrographic status epilepticus (E-SE) based on Salzburg and ACNS criteria,^22, 23^ interictal epileptiform discharges (IEDs) based on IFCN criteria,^24^ lateralized periodic discharges (LPDs), lateralized rhythmic delta activity (LRDA), and generalized periodic discharges (GPDs), following the American Clinical Neurophysiology Society nomenclature. ^23^ Focal and generalized slowing was also analyzed as a possible risk factor.

Further definitions and EEG parameter characterizations are provided in the Online Supplement.

### Design and Methodology

The flowchart in **Supplemental Figure 1** details the selection process for the study population. Initially, 1,177 post-stroke survivors from six centers were identified. Of these, six cohorts were included based on the availability of EEG data: three cohorts specifically designed to assess EEG after stroke and three cohorts that routinely acquired EEG data as part of standard clinical practice. Among these, 1,105 post-stroke survivors had EEG acquired within 7 days of stroke. This group was then divided into the derivation cohort (n=980) and the validation cohort (n=125). Within the derivation cohort, 336 post-stroke survivors had cEEG data available.

### Statistics

First, we used multivariable Cox proportional hazards regression in the derivation cohort to assess the relationship between clinical and electrographic findings and the time to post-stroke epilepsy while adjusting for known predictors of post-stroke epilepsy: National Institutes of Health Stroke Scale (NIHSS) score at admission, cortical involvement, involvement of the middle cerebral artery (MCA) territory, and large artery atherosclerosis.^4, 8^Cases were censored at the time of first RSyS or last follow-up. Adjusted risk estimates for post-stroke epilepsy were obtained from these multivariable Cox regression models.

Given that electrographic findings demonstrated a higher prognostic value in stroke survivors without ASyS, as described in the results, we developed a novel predictive model, called SeLECT-EEG, specifically for this subgroup to enhance the prediction of post-stroke epilepsy. Variables previously reported in the SeLECT_2.0_ prognostic model^4^ were retained in the new SeLECT-EEG model. This was done to maintain consistency with previous models and because these variables were repeatedly shown to be associated with post-stroke epilepsy in previous studies. We assigned integer values to the retained variables based on their adjusted hazard ratios (aHR, see **Table 1**). To ensure comparability between the new SeLECT-EEG prediction model and the previous SeLECT_2.0_ model and to adjust for the higher propensity of receiving an EEG in those at greater risk of post-stroke epilepsy - which may have artificially inflated risk estimates - we performed inverse probability weighting using 3,711 stroke survivors from the remaining cohorts in the SeLECT registry as a reference. Kaplan-Meier estimate plots were applied to compare the observed RSyS risk in those with vs. without significant electrographic findings from the initial Cox proportional hazards regression model.

**Table 1:** Comparison of the original SeLECT_2.0_ scoring system (in stroke survivors without acute-symptomatic seizures) and the modified SeLECT-EEG score including electrographic findings. To calculate an individual’s SeLECT-EEG score, the points associated with each predictor (aHR < 2.5 =1 point and aHR > 2.5 = 2 points) can be added to obtain the total risk score. The total score ranges from 0 to 8 points. IED, interictal epileptic activity; GPD, generalised periodic discharges; LPD, lateralized periodic discharges; LRDA, lateralized rhythmic delta activity; ESei, electrographic seizures; E-SE, electrographic status epilepticus

| Criteria | aHR<br>(95% CI) | SeLECT-EEG<br>(points) | SeLECT<sub>2.0</sub><br>(points) |
| --- | --- | --- | --- |
| **Severity of stroke** |  |  |  |
| NIHSS 4-10 | 1.1 (0.5-2.1) | 1 | 1 |
| NIHSS $\geq 11$ | 2.9 (1.6-5.3) | 2 | 2 |
| **Large-artery atherosclerosis** |  |  |  |
| No | - | 0 | 0 |
| Yes | 1.3 (0.7-2.1) | 1 | 1 |
| **Cortical involvement** |  |  |  |
| No | - | 0 | 0 |
| Yes | 2.5 (1.4-4.7) | 2 | 2 |
| **Territory of MCA involvement** |  |  |  |
| No | - | 0 | 0 |
| Yes | 1.3 (0.7-2.7) | 1 | 1 |
| **Electrographic Biomarkers** |  |  |  |
| Regional slowing |  |  |  |
| No | - | 0 | - |
| Yes | 2.4 (1.4-4.4) | 1 | - |
| All epileptiform activity (IED, GPD, LPD, LRDA, ESei, E-SE) |  |  |  |
| No | - | 0 | - |
| Yes | 2.0 (1.2-3.1) | 1 | - |
| **Maximum Points** |  | **8** | **6** |

To assess model performance, we calculated discrimination and calibration in the derivation cohort and in an independent external validation cohort after imputing for missing generalized and regional slowing values. To evaluate the novel model’s discrimination - the ability to distinguish between high and low-risk cases - we estimated the C statistic (95% CI). Recognizing that predictive models derived from multivariable regression may be optimistic and potentially overestimate predictions when applied to new cohorts, we introduced a shrinkage factor. This factor was estimated through 1000 bootstrapped random samples to adjust the C statistic for overoptimism, a technique previously employed to enhance model generalizability.^25, 26^ We also assessed model calibration - the agreement between predicted and observed risks - using calibration plots (see **Supplemental Figure 2**). A 45° diagonal line represents perfect calibration, whereas deviations above or below this line reflect under- or overprediction. We used a leave-one-cohort-out strategy to cross-validate model performance (see **Supplemental Figure 3**). Furthermore, mediation analysis was performed to assess the role of epileptiform activity as a mediator in the relationship between regional and generalized slowing and late seizure outcomes.

Finally, we calculated the Change of Occurrence of a Seizure in the next Year (COSY),^27^ relevant for assessing driving fitness in individuals with seizures, using standard conditional risk definitions (see **Supplemental Figure 4**).^28^ Suggested COSY thresholds are <20-40% for private driving and <2% for professional driving, though these may vary by local regulations.^29-31^

## Results

The baseline characteristics and frequency of EEG parameters for the derivation cohort are shown in **Table 2, Supplemental Table 1 and Supplemental Table 2**. In univariable analysis, epileptiform activity (including IED, LPD, GPD, LRDA, ESz, E-SE; RR 3.5, 95% CI: 1.5-5.2, p<0.001), regional slowing (HR 2.6, 95% CI: 1.7-4.0, p<0.001) and generalised slowing (HR 2.7, 95% CI: 1.9-4.0, p<0.001) were associated with post-stroke epilepsy **(Figure 1**). In multivariable analysis epileptiform activity (aHR 2.0, 95% CI 1.3-3.0, p=0.001) and regional slowing (aHR 1.9, 95% CI 1.2-2.9, p=0.004) were independently associated with post-stroke epilepsy **(Figure 2).** Generalized slowing, however, did not show a significant independent association in the multivariable analysis (**Supplemental Table 3**). Stroke survivors with epileptiform activity had a 42% risk (95% CI 30%-49%) of developing post-stroke epilepsy 5 years after stroke, compared to a 13% risk (95% CI 9%-16%) in those without (**Figure 2A**). Additionally, the 5-year risk of post-stroke epilepsy was twice as high in those with regional slowing (24%, 95% CI 18%-29%) compared to those without it (11%, 95% CI 5%-15%, **Figure 2B**).

**Figure 1:**
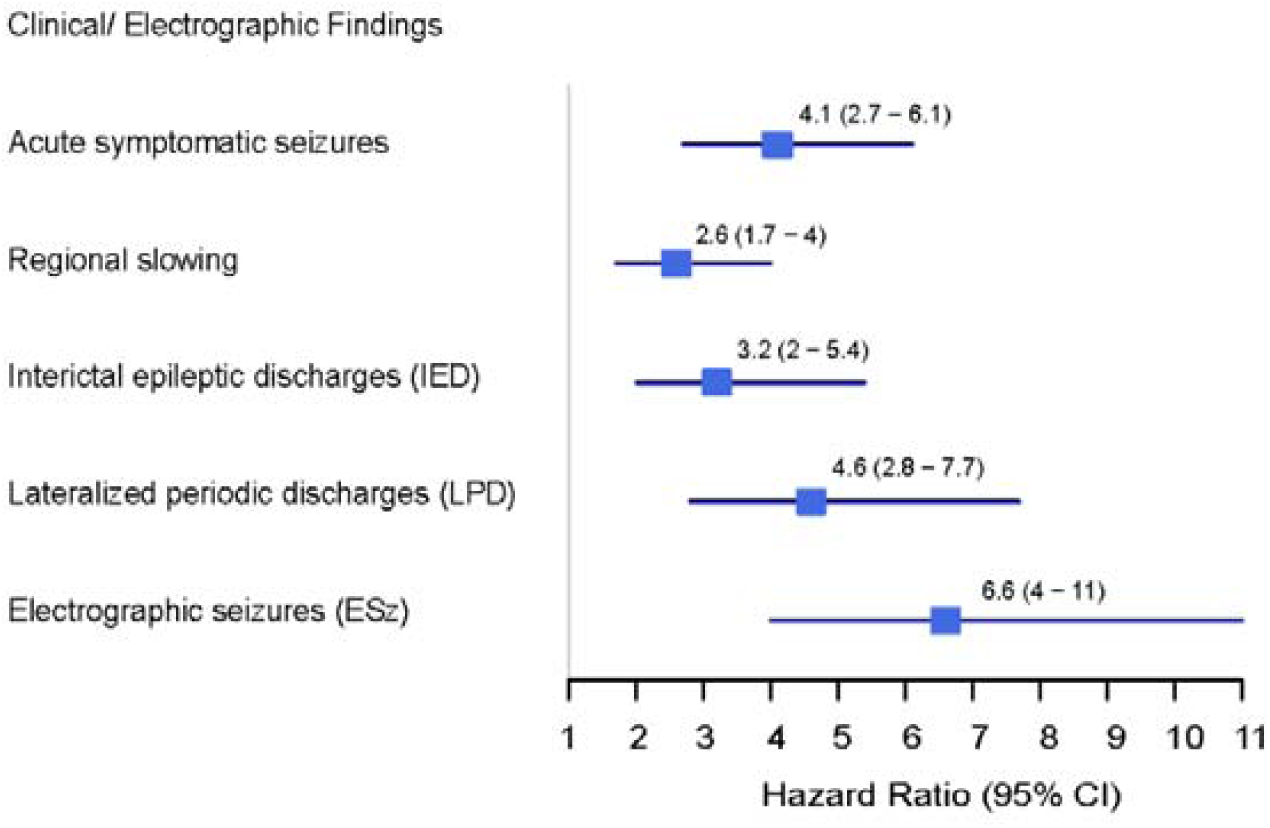
Forest Plot of Clinical and Electrographic Findings Associated with Post-Stroke Epilepsy Risk. This forest plot illustrates the HR and 95%-CI from **Table 1** for various clinical and electrographic findings in relation to the development of PSE. ASyS is shown alongside electrographic biomarkers such as regional slowing, IED, LPD, and ESz. Each of these variables significantly increases the risk of PSE, with ORs ranging from 2.6 to 6.6. Interestingly, ESz exhibited an HR comparable to clinical seizures (ASyS).

**Figure 2:**
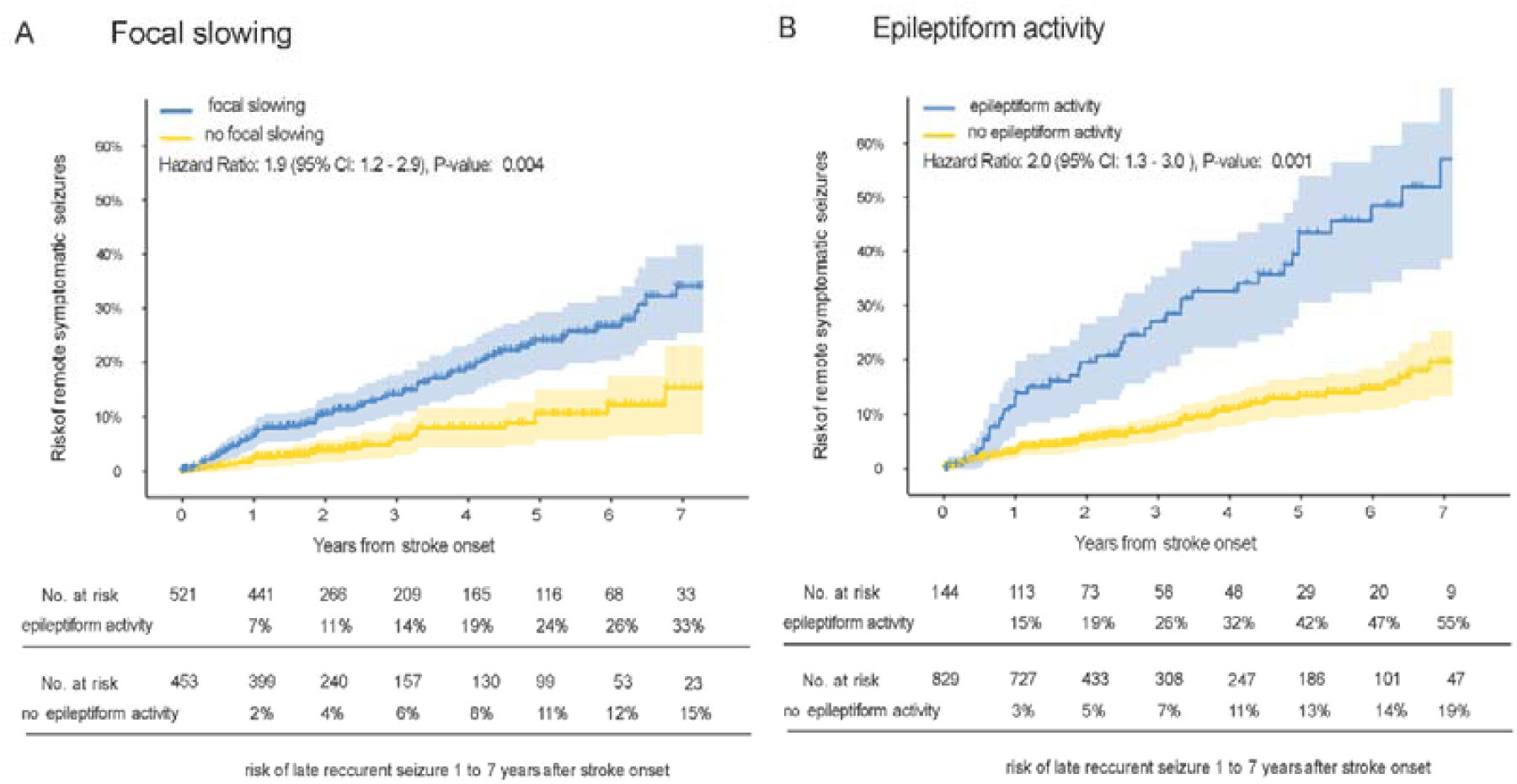
Risk of epilepsy in individuals with or without regional slowing or any epileptiform activity. Kaplan-Meier estimates (n = 980) for the time to post-stroke epilepsy are presented, comparing individuals with any epileptiform activity (Panel A) or regional slowing (Panel B). Separate survival curves are illustrated for those with (red) and without (blue) epileptiform activity (Panel A) or regional slowing (Panel B). Individuals exhibiting epileptiform activity or regional slowing demonstrated a significantly higher risk of developing post-stroke epilepsy, indicating a higher likelihood of remote symptomatic seizures, compared to those without these conditions (epileptiform activity: p < 0.001; regional slowing: p = 0.004).

**Table 2:**
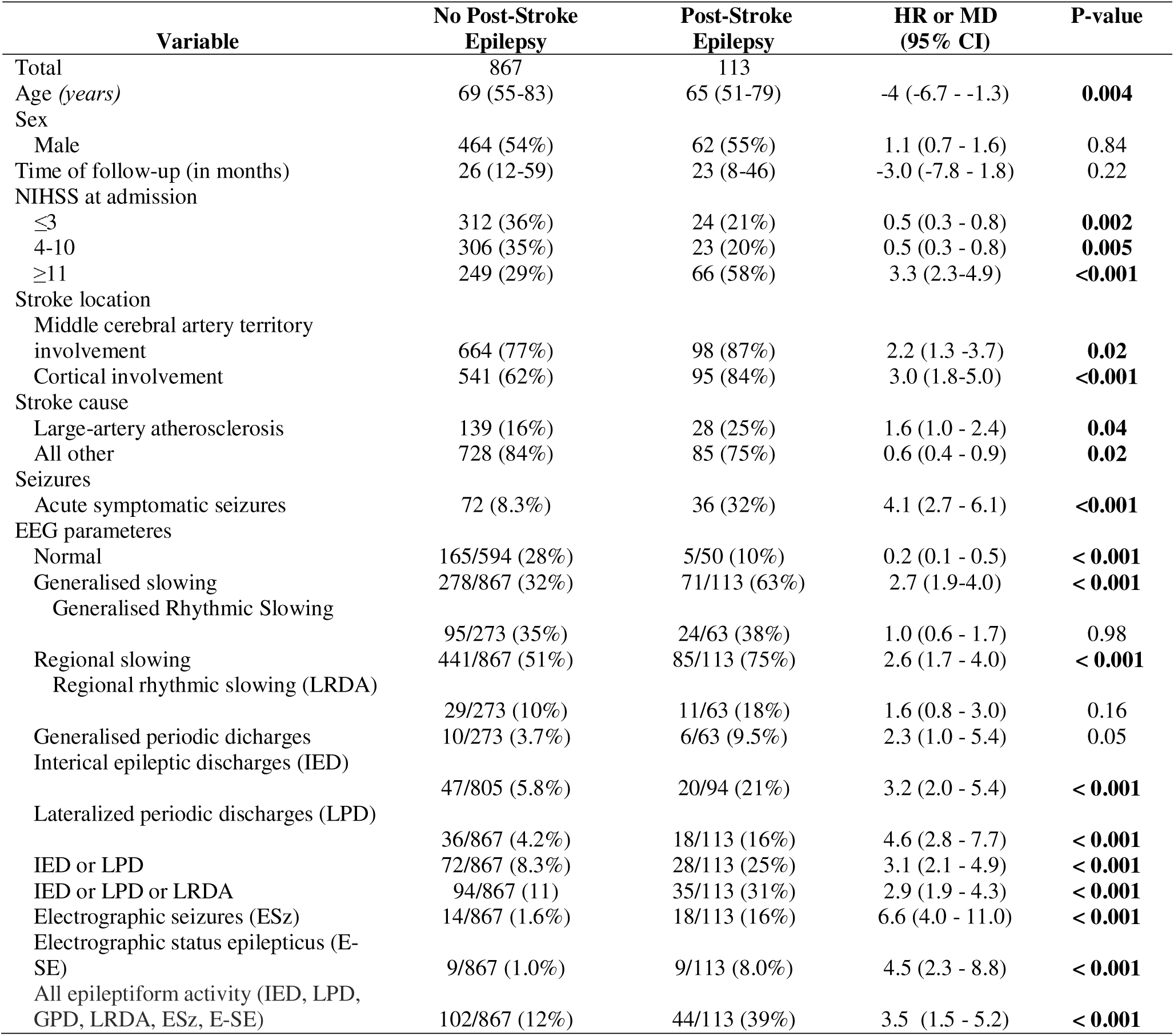
Patient and clinical characteristics, stratified by whether patient had a remote synptomatic seizure (> 7 days after stroke) in derivation cohorts (EEG variables including data from cEEG, n=980) This table presents the hazard ratios (HR) and mean differences (MD) comparing patients with post-stroke epilepsy to those without in the derivation cohort (n=980), calculated using univariate Cox regression. The HRs are derived from the perspective of PSE, emphasizing significant findings. Mean differences are provided for continuous variables such as age and follow-up duration. Notable significant findings include an increased risk for interictal epileptic discharges (IED) (HR 3.2, 95% CI: 2.0–5.4), lateralized periodic discharges (LPD) (HR 4.6, 95% CI: 2.8–7.7), and electrographic seizures (ESz) (HR 6.6, 95% CI: 4.0–11.0). Additionally, both generalized slowing (HR 2.7, 95% CI: 1.9–4.0) and regional slowing (HR 2.6, 95% CI: 1.7–4.0) are strongly associated with the PSE, indicating a strong link between these EEG abnormalities and PSE.

| Variable | No Post-Stroke Epilepsy | Post-Stroke Epilepsy | HR or MD (95% CI) | P-value |
| --- | --- | --- | --- | --- |
| Total | 867 | 113 |  |  |
| Age (years) | 69 (55-83) | 65 (51-79) | -4 (-6.7 - -1.3) | **0.004** |
| Sex |  |  |  |  |
| Male | 464 (54%) | 62 (55%) | 1.1 (0.7 - 1.6) | 0.84 |
| Time of follow-up (in months) | 26 (12-59) | 23 (8-46) | -3.0 (-7.8 - 1.8) | 0.22 |
| NIHSS at admission |  |  |  |  |
| ≤3 | 312 (36%) | 24 (21%) | 0.5 (0.3 - 0.8) | **0.002** |
| 4-10 | 306 (35%) | 23 (20%) | 0.5 (0.3 - 0.8) | **0.005** |
| ≥11 | 249 (29%) | 66 (58%) | 3.3 (2.3-4.9) | **<0.001** |
| Stroke location |  |  |  |  |
| Middle cerebral artery territory involvement | 664 (77%) | 98 (87%) | 2.2 (1.3 -3.7) | **0.02** |
| Cortical involvement | 541 (62%) | 95 (84%) | 3.0 (1.8-5.0) | **<0.001** |
| Stroke cause |  |  |  |  |
| Large-artery atherosclerosis | 139 (16%) | 28 (25%) | 1.6 (1.0 - 2.4) | **0.04** |
| All other | 728 (84%) | 85 (75%) | 0.6 (0.4 - 0.9) | **0.02** |
| Seizures |  |  |  |  |
| Acute symptomatic seizures | 72 (8.3%) | 36 (32%) | 4.1 (2.7 - 6.1) | **<0.001** |
| EEG parameteres |  |  |  |  |
| Normal | 165/594 (28%) | 5/50 (10%) | 0.2 (0.1 - 0.5) | **< 0.001** |
| Generalised slowing | 278/867 (32%) | 71/113 (63%) | 2.7 (1.9-4.0) | **< 0.001** |
| Generalised Rhythmic Slowing | 95/273 (35%) | 24/63 (38%) | 1.0 (0.6 - 1.7) | 0.98 |
| Regional slowing | 441/867 (51%) | 85/113 (75%) | 2.6 (1.7 - 4.0) | **< 0.001** |
| Regional rhythmic slowing (LRDA) | 29/273 (10%) | 11/63 (18%) | 1.6 (0.8 - 3.0) | 0.16 |
| Generalised periodic discharges | 10/273 (3.7%) | 6/63 (9.5%) | 2.3 (1.0 - 5.4) | 0.05 |
| Interictal epileptic discharges (IED) | 47/805 (5.8%) | 20/94 (21%) | 3.2 (2.0 - 5.4) | **< 0.001** |
| Lateralized periodic discharges (LPD) | 36/867 (4.2%) | 18/113 (16%) | 4.6 (2.8 - 7.7) | **< 0.001** |
| IED or LPD | 72/867 (8.3%) | 28/113 (25%) | 3.1 (2.1 - 4.9) | **< 0.001** |
| IED or LPD or LRDA | 94/867 (11) | 35/113 (31%) | 2.9 (1.9 - 4.3) | **< 0.001** |
| Electrographic seizures (ESz) | 14/867 (1.6%) | 18/113 (16%) | 6.6 (4.0 - 11.0) | **< 0.001** |
| Electrographic status epilepticus (E-SE) | 9/867 (1.0%) | 9/113 (8.0%) | 4.5 (2.3 - 8.8) | **< 0.001** |
| All epileptiform activity (IED, LPD, GPD, LRDA, ESz, E-SE) | 102/867 (12%) | 44/113 (39%) | 3.5 (1.5 - 5.2) | **< 0.001** |

### Prognostic modeling in stroke-survivors without ASyS

Besides already known predictors of post-stroke epilepsy, we performed a multivariate Cox-regression model in stroke survivors (**Supplemental Table 4**) to find new electrographic biomarkers. Variables retained in the final model were stroke severity at admission (NIHSS), stroke location with middle cerebral artery territory and cortical involvement, large-vessel atherosclerotic stroke etiology, epileptiform activity, and regional slowing. The calculation of the new model termed SeLECT-EEG and ranging from 0 to 8 points, is shown in **Table 1**.

The full results of the multivariable model and other variables independently associated with post-stroke epilepsy (stroke severity and location) in stroke survivors are shown in **Supplemental Table 4.**

The SeLECT-EEG model demonstrated better discrimination for time to post-stroke epilepsy compared to the original SeLECT_2.0_ model in stroke survivors without ASyS (C-Statistic 0.75 [95%-CI 0.71-0.80] vs. 0.71 [95%-CI 0.65-0.76], p < 0.001; **Supplemental Table 5**). For stroke survivors with ASyS, the C-Statistics were not significantly different between the SeLECT_2.0_ and SeLECT-EEG models (both 0.59, p = 0.97; **Supplemental Table 5 and Supplemental Table 6**). Due to these findings and the low number of late seizure events in the ASyS group, we developed the model only in stroke survivors without ASyS. SeLECT-EEG demonstrated a near-optimal calibration for outcomes although lacking data for lower probability outcomes (**Supplemental Figure 2**).

We cross-validated the results using a leave-one-cohort-out strategy (**Supplemental Table 7**). The lowest score SeLECT-EEG value (0 points) predicts a 2.5% risk (95% CI 0.5-4.4%) of post-stroke epilepsy 10 years following stroke, compared with a 95% risk (95% CI 67-99%) for the highest value (8 points, **Figure 3** and **Supplemental Figure 5** (comparison with SeLECT_2.0_ model)). In addition, we updated the values for COSY (**Supplemental Figure 4**), a parameter that may be helpful for assessing the risks of safe driving.^28^ The new SeLECT-EEG model was implemented in the “SeLECT score” smartphone applications available for iOS and Android and in the web-based calculator.

**Figure 3:**
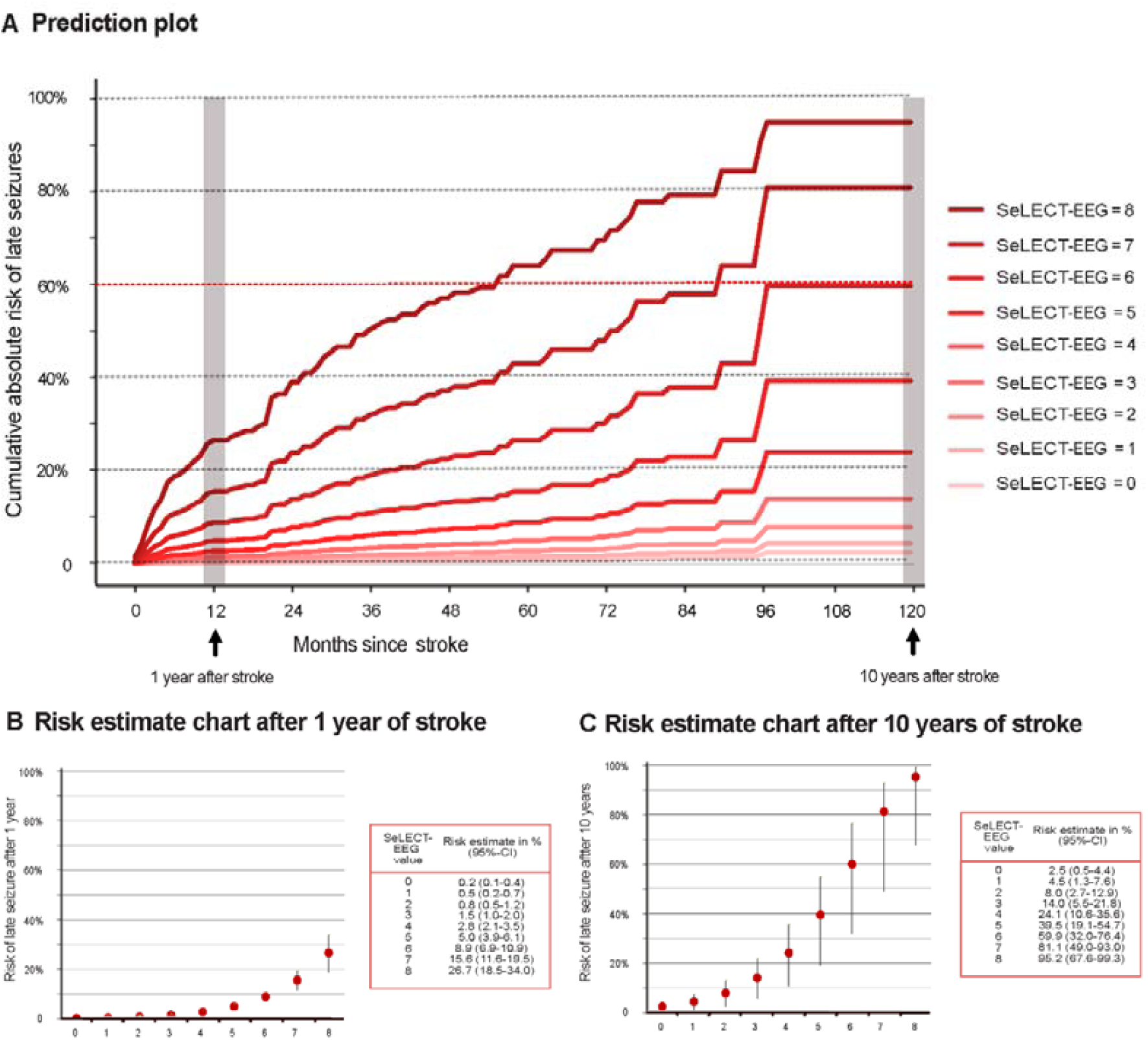
Predicted Risk of Post-Stroke Epilepsy according to a new prognostic model in stroke survivors without acute symptomatic seizures including electrographic findings Panel. **A** illustrates the predicted risk of unprovoked remote symptomatic seizures occurring 0-120 months post-stroke. The red curves represent different SeLECT-EEG values for stroke survivors including electrographic findings. Each curve corresponds to SeLECT-EEG values ranging from 0-8 (in red). **Panels B and C** present risk estimate charts for late seizures at 1 year and 10 years post-stroke, respectively, based on the SeLECT-ASyS. Vertical lines denote the 95% confidence intervals.

### Length of EEG recording

Longer EEG recordings had a higher yield of electrographic findings (**Supplemental Table 8 and Supplemental Figure 3**). Overall, some findings, such as normal patterns (short vs. cEEG counts: 207/199, 104%) and generalised (318/348, 91%) and regional (470/526, 89%) slowing, had high detection rates in short EEGs, comparable to cEEGs. However, other findings, such as epileptiform activity (105/146, 72%), particularly IED (44/67, 66%) and rhythmic patterns (e.g., LRDA; 16/40, 40%), showed lower detection rates in short EEGs. The increased detection rate of regional slowing and epileptiform activity enhanced the discrimination of the SeLECT-EEG model (c-statistics for short vs. continuous recordings: 0.72 [95%-CI 0.66-0.77] vs. 0.75 [95%-CI 0.71-0.80], p < 0.001, **Supplemental Table 6**).

Mediation analysis showed that epileptiform activity accounted for 30% of the total effect of both regional and generalized slowing on late seizure outcomes (**Supplemental Table 9**).

Furthermore, generalized slowing was more prevalent in severely affected stroke survivors (NIHSS ≥11) undergoing cEEG recordings (**Supplemental Table 8**).

### Case Example

As an example, we considered a 65-year-old male patient admitted to the hospital with an acute ischemic stroke of cardioembolic etiology, affecting the medial cerebral artery territory and involving the cortex, with 16 points on the NIHSS at admission. On day 2 after the stroke, a routine EEG was performed, which showed IEDs. The SeLECT_2.0_ score for this case is 5 points, predicting a 30% risk (95% CI 14-44%) of post-stroke epilepsy within 10 years after stroke. In contrast, the new model developed specifically for those without acute symptomatic seizures and available EEG data (SeLECT-EEG, **Table 1**) is 7 points, predicting 81% (95% CI 49-93%) risk of post-stroke epilepsy within 10 years.

## Discussion

In this study, we examined the predictive value of electrographic biomarkers detected on EEG within the first 7 days after ischemic stroke for post-stroke epilepsy. Our results show that specific EEG abnormalities, such as regional slowing and epileptiform activity (including IED, LPD, GPD, LRDA, Esz, and E-SE), are significantly associated with an increased risk of post-stroke epilepsy, particularly in stroke survivors without ASyS.

The question of how EEG findings can serve as predictors of post-stroke epilepsy is rooted in their ability to detect early signs of cortical hyperexcitability, a key factor in epileptogenesis.^32-34^ Electrographic seizures (ESz) and rhythmic or periodic patterns (RPPs), which are observed in as many as 15% of acute ischemic stroke survivors, are not merely markers of current neural dysfunction but also indicators of underlying processes that may lead to epilepsy.^10-13^ These EEG abnormalities share risk factors with post-stroke epilepsy, such as cortical involvement and stroke severity, suggesting that they can act as early warning signs of future seizures.^14^

Furthermore, EEG sensitivity to ischemic cortical injury enables it to detect subtle, preclinical changes that contribute to the development of epilepsy. This is particularly important because such changes might not yet be apparent through clinical symptoms alone. The predictive value of EEG is also supported by similar findings in TBI, where EEG abnormalities like IEDs and RPPs are consistently linked to an increased risk of post-traumatic epilepsy.^35-37^ These insights underscore the importance of incorporating EEG findings into seizure prediction models, as they provide a critical layer of prognostic information that complements clinical risk factors.

Building on this understanding, we developed the SeLECT-EEG model specifically for stroke survivors without ASyS. This model not only accurately captures the heightened risk of post-stroke epilepsy in this subgroup but also significantly outperforms the SeLECT_2.0_ model (C-Statistic 0.75 [95%-CI 0.71-0.80] vs. 0.71 [95%-CI 0.65-0.76], p < 0.001; **Supplemental Table 6**). In contrast, for stroke survivors with ASyS, the predictive accuracy of the SeLECT_2.0_ and SeLECT-EEG models did not differ significantly (both C-Statistics 0.59, p = 0.97). Given these findings and the likely distinct pathophysiological mechanisms in stroke survivors with ASyS (Schubert et al. 2024, under review), we focused the SeLECT-EEG model on those without ASyS.

Translating these findings into clinical practice, the SeLECT-EEG model emerges as the preferred tool for prognostication in stroke survivors without ASyS. The case example presented illustrates the notable differences in risk estimation when using SeLECT-EEG compared to SeLECT_2.0_, which could be clinically relevant and may influence treatment decisions and follow-up strategies. The model risk estimates offer a more nuanced assessment of post-stroke epilepsy risk, supported by its favorable calibration (**Supplemental Figure 2**).

In addition to these findings, cEEG monitoring demonstrated a higher detection rate of epileptiform activity compared to short EEG recordings, significantly improving the predictive accuracy of the SeLECT-EEG model (C-statistics: 0.75 vs. 0.72, p < 0.001; **Supplemental Table 6**). While short EEGs effectively detected generalized and regional slowing (**Supplemental Table 8-10,** and **Supplemental Figure 3**), they performed poorly in stroke survivors with higher NIHSS scores (Group 2 with cEGG, **Supplemental Table 8)**, where generalized slowing was more prevalent. This generalized slowing likely complicated the detection of focal slowing, while the short duration of the recording made it more challenging to detect epileptiform activity, underscoring the importance of continuous monitoring for identifying critical findings such as IEDs (**Supplemental Figure 3**).^18, 19^ Mediation analysis demonstrated that epileptiform activity accounted for around 30% of the effect of both regional and generalized slowing on late seizure outcomes, indicating that epileptiform activity is unlikely without the presence of either focal or generalized slowing (**Supplemental Table 9**). However, the optimal length of EEG monitoring remains unanswered, as cEEG offers benefits in certain high-risk subgroups, while normal short EEGs show high specificity in identifying stroke survivors unlikely to develop post-stroke epilepsy, reducing the need for extended monitoring (**Supplemental Table 8** and **Supplemental Figure 3**).

A key finding of this study is that stroke survivors without ASyS who have SeLECT-EEG scores of 7 or more face a greater than 60% 10-year risk of developing epilepsy. This risk level aligns with the ILAE’s definition of epilepsy.^20^ Although this definition traditionally requires an unprovoked seizure, some clinicians might consider preemptive treatment or extended follow-up, including EEGs for these high-risk stroke survivors, even in the absence of a seizure.^5, 38^ However, the efficacy of such prophylactic treatment remains uncertain.

In a previous smaller analysis of the SeLECT registry,^39^ we did not find significant associations between EEG findings and post-stroke epilepsy, likely due to the limited sample size and individual analysis of electrographic biomarkers. However, by expanding the dataset to include additional cohorts^18, 19^ and refining our approach to combine biomarkers and focus on stroke survivors without acute symptomatic seizures, we now demonstrate significant associations.

Our study has several strengths. To our knowledge, this is the largest multicenter cohort study of post-stroke seizures assessing the predictive power of electrographic biomarkers. We translated our findings into a clinical model and user-friendly prognostic tool accessible via both smartphone and web applications. The novel model outperforms the current state-of-the-art model.

However, our study has several limitations. First, the indication for EEGs and their analysis varied across the eleven international cohorts included in this study, spanning from 2002 to 2022. In some earlier cohorts, primarily those performing short EEGs (Group 1), the detection of certain rhythmic and periodic patterns may have been limited due to retrospective EEG assessments and differing criteria definitions. To correct for these variations and ensure comparability between the new SeLECT-EEG prediction model and the previous SeLECT2.0 model, inverse probability weighting was applied using data from the broader SeLECT cohort (n = 3,711).

Second, we grouped various EEG abnormalities, which may have obscured more specific prognostic factors. For example, generalized slowing was not consistently defined by a specific frequency, potentially leading to interpretational differences between cohorts.

Third, the SeLECT-EEG model was developed specifically for stroke survivors without ASyS because the event number in stroke survivors with ASyS was too low to improve upon the existing SeLECT_2.0_ prognostic model. Additionally, the C-statistics for the SeLECT-EEG and SeLECT_2.0_ models in the ASyS group were not significantly different from each other, and some clinical parameters necessary to adapt the model to the recently published SeLECT-ASyS framework (Schubert et. al 2024, under review) were missing.

Fourth, the overall post-stroke epilepsy rate in our study was relatively high at 11%, likely influenced by the stringent selection criteria for cEEG monitoring in certain cohorts, particularly those from Belgium^18^ and the USA^19^ (**Supplemental Table 7**). We also used inverse probability weighting to account for these differences and ensure the new SeLECT-EEG model was comparable to the SeLECT_2.0_ model.

Lastly, in the external validation cohort, there was missing data on regional and generalized slowing. We addressed this by using multiple imputations to handle the missing data, allowing for a more accurate validation of the model. While calibration was strong, and discrimination showed a trend towards better performance, though this was not statistically significant (**Supplemental Figure 2**, p = 0.32), likely due to the small sample size in this subgroup (N=119).

## Conclusions

Our study demonstrates that electrographic abnormalities detected on EEG during the acute phase of ischemic stroke, particularly regional slowing and epileptiform activity, are significant predictors of post-stroke epilepsy. The SeLECT-EEG model, developed specifically for stroke survivors without ASyS, outperforms the existing SeLECT_2.0_ model in predicting post-stroke epilepsy risk. CEEG monitoring, especially in selected stroke survivors with higher NIHSS scores and more severe generalized slowing, further enhances the detection of critical abnormalities, improving the model’s predictive accuracy.

Notably, certain SeLECT-EEG risk scores exceed a 60% 10-year risk of developing epilepsy, aligning with the ILAE’s definition of epilepsy.^20^ This finding suggests the potential for preemptive treatment or extended follow-up for high-risk stroke survivors, although the effectiveness of such approaches remains uncertain. Integrating these EEG-based insights into clinical practice could refine risk stratification in stroke survivors and help identify enriched cohorts for studies focused on preventive strategies for the development of post-stroke epilepsy.

## Authors Contributions

KMS, CB and MG conceptualized and designed the study. KMS, VD, ALO, CT, GN, NG, AS, MG, VP, CB contributed to the acquisition and analysis of the data. KMS, CB, VP, NG and MG contributed to the interpretation of the data. KMS and MG drafted the manuscript and figures. All co-authors revised the manuscript for intellectual content.

## Data Availability

All data produced in the present study are available upon reasonable request to the authors

