## Supplemental for "The role of EEG in predicting post-stroke seizures and an updated prognostic model (SeLECT-EEG)"

-- ONLINE SUPPLEMENT --

**Content:**

### Supplemental Methods: Description of Cohorts

The full cohort (n=4888) consisted of eleven international subcohorts participating in a registry assessing post-stroke seizures incepted as part of the SeLECT study. Six cohorts had EEG data available. The description of the cohorts of the original SeLECT study can be found elsewhere (Schubert 2024). Three out of these subcohorts (Portugal, Switzerland (1), Switzerland (2)) were part of the original SeLECT study, and three (Germany, USA and Belgium), were included additionally.

**Portugal**

The Portuguese cohort consisted of 147 adults admitted to a university hospital in Lisbon for neuroimaging-confirmed anterior circulation ischemic stroke. This prospective, longitudinal study spanned 24 months, with an additional 12-month follow-up period. Eligibility criteria included prior independence (mRS ≤1), a NIHSS score of at least 4 upon emergency department admission, an acute ischemic brain lesion in the anterior circulation, and no history of epileptic seizures, significant head trauma, or brain surgery. Standardized clinical and diagnostic assessments were conducted during admission and at discharge. Follow-up for epileptic seizures and functional outcomes was performed via blinded phone interviews at six months and clinical appointments at twelve months post-stroke. EEG monitoring was conducted within the first 72 hours after stroke using a 64-channel video-EEG for up to 60 minutes, including eyes closed resting, eyes open, hyperventilation, and photic stimulation maneuvers. EEG analysis focused on detecting background activity slowing (Noachtar 2017), asymmetry (Hirsch 2013), suppression patterns (regional, hemispheric, or generalised), nonrhythmic slow wave activity (NRSA, Noachtar 2017), and rhythmic slow-wave activity (RSA), including lateralized rhythmic delta activity (LRDA, Hirsch 2013). Other EEG features monitored were interictal epileptiform activity, lateralized periodic discharges, and electrographic seizures. The EEGs were reviewed and classified by a certified clinical neurophysiologist using international criteria (Nocahtar 2017; Hirsch 2013, Beniczky 2013), with consensus reached on any uncertainties. Neuroimaging studies were analyzed by a senior neuroradiologist using the Alberta Stroke Program Early CT Score (ASPECTS) to quantify MCA infarct size and location. The primary outcome was the occurrence of at least one remote symptomatic seizure within the first year post-stroke.

**Switzerland (1)**

The Swiss cohort (1) included consecutive individuals aged 18 years or older who were admitted with a first-ever, neuroimaging-confirmed ischemic stroke to a tertiary referral center in St. Gallen between January 1, 2002, and December 31, 2008. The study excluded individuals with transient ischemic attacks (n=495), a previous history of stroke (n=250), primary hemorrhagic stroke (n=94), previous history of seizures (n=43), re-infarction during follow-up (n=9), and those with potentially epileptogenic comorbidities such as alcohol or drug abuse (n=60), intracranial tumors (n=28), cerebral venous thrombosis (n=11), severe traumatic brain injury (n=12), brain surgery (n=4), or other conditions like cerebral arteriovenous malformations, large cerebral aneurysms, cerebral vasculitis, hydrocephalus, and cerebral abnormalities of undetermined etiology (n=15). A total of 85 participants were lost to follow-up or died before the follow-up could be conducted.

A neurologist analyzed baseline characteristics at admission, and a diagnosis of stroke was confirmed at discharge. Brain scans were performed using the best available imaging modality, with MRI used in 954 (80%) of 1200 participants and CT in 246 (21%). All participants with EEG (final n=330) were followed up after a median of 28 months (IQR 21–47) with a structured telephone interview based on a validated questionnaire to detect seizures. For participants unable to complete the questionnaire, close relatives, nursing staff, or their general practitioner were interviewed. Positive responses led to a face-to-face neurological consultation to determine the epileptic nature of these episodes and to exclude seizure mimics. If the neurologist suspected a cause of epileptic seizures other than the index ischemic stroke, follow-up imaging was requested to rule out a co-pathology or re-infarction. The same EEG criteria used in the Portuguese cohort were employed to evaluate the participants. These criteria included monitoring for background activity slowing, asymmetry, suppression patterns, nonrhythmic slow wave activity (NRSA) characterized as continuous or intermittent slow activity without a constant period in specific brain regions, and rhythmic slow-wave activity (RSA) including definitions like lateralized rhythmic delta activity (LRDA). Additional EEG features such as interictal epileptiform activity, lateralized periodic discharges, and electrographic seizures were also scrutinized.

**Switzerland (2)**

The Swiss cohort (2) was part of the Biomarker Signature of Stroke Aetiology (BIOSIGNAL) study, a prospective, observational, multicenter, inception cohort study focused on evaluating and validating selected blood biomarkers for acute ischemic stroke. The participants, all older than 18 years, were recruited at the University Hospital Zurich, Switzerland, from October 2014 to April 2022, upon giving informed consent. Exclusions included individuals with transient ischemic attacks, intracranial tumors (n=17), severe traumatic brain injury (n=3), history of brain surgery (n=7), or other potentially epileptogenic comorbidities such as cerebral arterio-venous malformations, large cerebral aneurysms, cerebral vasculitis, hydrocephalus, and cerebral abnormalities of undetermined etiology (n=25). Participants with a history of seizures prior to stroke or cerebral venous thrombosis were also excluded. Out of the initial group, three participants were lost to follow-up, and 94 died before subsequent evaluations could be conducted, leaving a cohort of 1016 participants. Acute symptomatic seizures within the first 7 days post-stroke were documented for each participant. 227 stroke survivors had EEG data available. Standardized follow-ups at 3 and 12 months involved either an outpatient visit or a structured telephone interview by a neurologist to ascertain any unprovoked remote symptomatic seizures. Long-term follow-up, conducted 60 months post-stroke, was performed using chart review.

For the EEG assessment, similar criteria to those used in the Portuguese cohort were applied to monitor neurological activity and potential complications post-stroke. This included detecting background activity slowing, asymmetry, and various suppression patterns. Both nonrhythmic slow wave activity (NRSA), characterized as continuous or intermittent slow activity without a constant period, and rhythmic slow-wave activity (RSA) were monitored. Specific patterns such as lateralized rhythmic delta activity (LRDA) and other features including interictal epileptiform activities, lateralized periodic discharges, and electrographic seizures were also evaluated.

**Germany**

The German cohort prospectively enrolled all stroke survivors aged 18 years or older who were consecutively admitted to the Department of Neurology at Philipps-University Marburg, Germany, with a diagnosis of ischemic stroke over a 5-month period. Stroke survivors were screened for seizures during hospital treatment and at routine follow-ups at 6 and 12 months post-stroke. Exclusion criteria included stroke due to subarachnoid hemorrhage, cerebral venous sinus thrombosis, established epilepsy, and insufficient data. Demographic, clinical, radiological, laboratory, neurophysiologic, and outcome data, including the modified Rankin Scale (mRS), were collected at hospital admission. All stroke survivors underwent brain imaging (CT, MRI, or both) to determine stroke type, with results reviewed by a neuroradiologist. EEGs were performed at the attending physician’s discretion during hospital admission and were independent of the study protocol. Standard EEGs of at least 23 minutes were recorded using the international 10–20 electrode placement system. EEG recordings were reviewed by two epileptologists who were blinded to the clinical outcomes. In cases of disagreement, the EEGs were re-evaluated until a consensus was reached. Seizures and epilepsy were classified according to the International League Against Epilepsy (ILAE) classification.

**Belgium**

The Belgium cohort involved a retrospective analysis (n=81 stroke survivors) from the prospective continuous EEG (cEEG) and stroke registries at the Hôpital Universitaire de Bruxelles—Hôpital Erasme in Brussels, Belgium. The study included all consecutive adult stroke survivors (≥18 years) who underwent cEEG during the acute ischemic stroke phase between January 1, 2015, and December 31, 2019. cEEG was routinely performed for stroke survivors with non-lacunar supratentorial stroke with an NIHSS score >8, early clinical seizures, or unexplained early neurological deterioration. Stroke survivors with less than one year of follow-up after stroke, prior epilepsy, or epilepsy possibly due to another cause were excluded. The primary outcome of the study was post-stroke epilepsy (PSE), defined as the occurrence of at least one spontaneous seizure more than 7 days after stroke onset.

Clinical and electrographic data collected included demographics, early clinical seizures, stroke etiology (according to the TOAST classification), admission NIHSS score, stroke territory, and presence of cortical involvement on imaging. Additional data included acute phase treatment and follow-up duration. Outcomes measured were the occurrence of PSE and the modified Rankin Scale scores at the last follow-up, typically obtained during neurovascular, epilepsy, geriatrics, or general neurology follow-up visits.

For EEG analysis, one author, blinded to clinical data and primary outcomes, reviewed the EEG recordings for seizures (either electrographic or electroclinical), highly epileptogenic RPPs (including brief ictal rhythmic discharges, lateralized periodic discharges, bilateral independent periodic discharges and lateralized rhythmic delta activity, Hirsch 2022). Antiseizure medication was initiated in most stroke survivors with acute symptomatic clinical seizures and tapered during outpatient follow-up in the absence of seizure recurrence.

**USA**

The USA cohort (n=279) was formed utilizing a prospectively maintained stroke and EEG database to identify adults who presented with acute ischemic stroke (AIS) from April 1, 2012, to March 31, 2018. Stroke survivors underwent cEEG monitoring within 7 days of the last known well time, typically indicated for unexplained altered mental status or motor events suggestive of seizure. Each cEEG session began with a preliminary 20-minute EEG screening. The cEEG data and associated clinical records were reviewed by a research associate (LE), who was blinded to the data, to ascertain cases of post-stroke epilepsy (PSE) and matched controls. Stroke survivors with a history of epilepsy prior to AIS were excluded. Those who experienced clinical seizures after hospital discharge, as recorded by their treating physicians, were identified as having PSE.

The electronic medical records (EMR) were subsequently reviewed to collect acute clinical, neuroimaging, EEG, and anti-seizure medication data. The EEG analysis included looking for epileptiform abnormalities (EAs), such as electrographic seizures based on the Salzburg criteria (Beniczky 2013), isolated sharp waves (SWs), lateralized periodic discharges (LPDs), lateralized rhythmic delta activity (LRDA), and generalised periodic discharges (GPDs), classified according to the American Clinical Neurophysiology Society nomenclature (Hirsch 2022).

### Supplemental Methods: Informed consent procedures

All subjects in the Swiss (2) and Portuguese cohort and those having a face-to-face interview in the Swiss (1) cohort gave written informed consent. All subjects evaluated by telephone in the Swiss (1) cohort gave verbal informed consent. According to Swiss law the regional ethical committees exempted these cohorts from requiring written informed consent. The USA cohort was formed under IRB-approved protocols. The study from the belgian cohort was approved by the Erasme Hospital Ethics Committee, which waived the need for informed consent.

### Supplemental Methods: Other definitions

We used definitions and classifications of the International League Against Epilepsy (ILAE) for seizures types and epilepsy^1-3^ and the World Health Organization for stroke.^4^

Stroke severity was measured with the National Institutes of Health Stroke Scale (NIHSS). Stroke etiology was classified according to the Trial of Org 10172 in Acute Stroke Treatment (TOAST) classification.^5^

Supplemental Table 1: *Baseline characteristics of cohorts (n=1177)*

| **Variable** | **N (%) or**  **median (IQR)** |
| --- | --- |
| Cohort |  |
| Portugal | 147 (12%) |
| Switzerland (1) | 330 (28%) |
| Switzerland (2) | 227 (19%) |
| Belgium | 81 (7%) |
| USA | 267 (23%) |
| Germany (1) | 125 (11%) |
| Age *(years)* | 71 (60-79) |
| Sex |  |
| Male | 631 (54%) |
| Female | 541 (46%) |
| Time of follow-up (in months) | 23 (12-53) |
| NIHSS at admission |  |
| ≤3 | 410 (35%) |
| 4-10 | 413 (35%) |
| ≥11 | 354 (30%) |
| Stroke location |  |
| Middle cerebral artery territory involvement | 890 (76%) |
| Cortical involvement | 745 (63%) |
| Stroke cause |  |
| Large-artery atherosclerosis | 225 (19%) |
| All other | 952 (81%) |
| Clinical seizures |  |
| Early seizures | 116 (10%) |
| Late seizures | 138 (12%) |
| EEG parameteres |  |
| Normal | 199/980 (20%) |
| Generalised slowing | 349/980 (36%) |
| Generalised Rhythmic Slowing | 199/336 (35%) |
| Regional slowing | 526/980 (54%) |
| Regional rhythmic slowing (LRDA) | 40/336 (12%) |
| Generalised periodic discharges (GPDs) | 16/336 (4.8%) |
| Sporadic interictal epileptic activity (IEA) | 80/1024 (7.8%) |
| Lateralized periodic discharges (LPDs) | 54/980 (5.5%) |
| IEA or LPDs | 100/980 (10%) |
| IEA or LPDs or LRDA | 129/980 (13%) |
| Electrographic seizures (ESz) | 32/980 (3.3%) |
| Electrographic status epilepticus (ESE) | 18/980 (1.8%) |
| All epileptiform activities (IEA, GPDs, LPDs, LRDA, ESz, ESE) | 146/980 (15%) |

NIHSS, National Institutes of Healthy Stroke Scale.

Supplemental Table 2: *Baseline characteristics of derivation cohorts with available EEG (n=980)*

| **Variable** | **N (%) or**  **median (IQR)** |
| --- | --- |
| Cohort |  |
| Portugal | 147 (15%) |
| Switzerland (1) | 270 (28%) |
| Switzerland (2) | 227 (23%) |
| Belgium | 81 (8%) |
| USA | 255 (26%) |
| Age *(years)* | 71 (60-79) |
| Sex |  |
| Male | 526 (54%) |
| Female | 454 (46%) |
| Time of follow-up (in months) | 25 (12-57) |
| NIHSS at admission |  |
| ≤3 | 336 (34%) |
| 4-10 | 329 (34%) |
| ≥11 | 315 (32%) |
| Stroke location |  |
| Middle cerebral artery territory involvement | 762 (78%) |
| Cortical involvement | 636 (65%) |
| Stroke cause |  |
| Large-artery atherosclerosis | 167 (17%) |
| All other | 813 (83%) |
| Clinical seizures |  |
| Early seizures | 108 (11%) |
| Late seizures | 113 (12%) |
| EEG parameters |  |
| Normal | 199/980 (20%) |
| Generalised Slowing | 349/980 (36%) |
| Generalised Rhythmic Slowing | 199/336 (35%) |
| Regional slowing | 526/980 (54%) |
| Regional rhythmic slowing (LRDA) | 40/336 (12%) |
| Generalised periodic discharges | 16/336 (4.8%) |
| Sporadic interictal epileptic activity (IEA) | 67/899 (7.5%) |
| Lateralized periodic discharges (LPDs) | 54/980 (5.5%) |
| IEA or LPD | 100/980 (10%) |
| IEA or LPDs or LRDA | 129/980 (13%) |
| Electrographic seizures (ESz) | 32/980 (3.3%) |
| Electrographic status epilepticus (ESE) | 18/980 (1.8%) |
| All epileptiform activities (IEA, GPDs, LPDs, LRDA, ESz, ESE) | 146/980 (15%) |

NIHSS, National Institutes of Healthy Stroke Scale.

Supplemental Table 3: *Results of multivariate Cox- regression models of time to first remote symptomatic seizure (n=980 )in derivation cohorts*

|  | **Cox-regression** | |
| --- | --- | --- |
| **Variable** | **aHR**  **(95% CI)** | **P value** |
| Stroke severity at admission |  |  |
| NIHSS 4-10 | 0.8 (0.4-1.4) | 0.42 |
| NIHSS ≥11 | 1.9 (1.1-3.1) | **0.019** |
| Stroke location |  |  |
| Middle cerebral artery territory involvement | 1.5 (0.9-2.6) | 0.16 |
| Cortical involvement | 2.0 (1.2-3.3) | **0.011** |
| Large-artery atherosclerosis (LAA) | 1.4 (0.9-2.2) | 0.11 |
| Early seizure | 2.8 (1.9-4.2) | **< 0.001** |
| EEG parameters |  |  |
| Generalised slowing | 1.4 (0.9-2.2) | 0.14 |
| Regional slowing | 1.9 (1.2-2.9) | **0.004** |
| All epileptiform activities (IEA, GPDs, LPDs, LRDA, ESz, ESE) | 2.0 (1.3-3.0) | **0.001** |

IEA, sporadic interictal epileptic activity; GPDs, generalised periodic discharges; LPDs, lateralized periodic discharges; LRDA, lateralized rhythmic delta activity; ESz, electrographic seizures; ESE, electrographic status epilepticus. Early seizure are clinical early seizures.

Supplemental Table 4: *Results of multivariate Cox- regression models of time to first remote symptomatic seizure in stroke survivors without acute-symptomatic seizures (n=872) in derivation cohorts*

|  | **Cox-regression**  **(full model)** | |
| --- | --- | --- |
| **Variable** | **aHR**  **(95% CI)** | **P value** |
| Stroke severity at admission |  |  |
| NIHSS 4-10 | 1.0 (0.5-2.0) | 0.94 |
| NIHSS ≥11 | 2.4 (1.3-4.6) | **0.006** |
| Stroke location |  |  |
| Middle cerebral artery territory involvement | 1.3 (0.6-2.7) | 0.48 |
| Cortical involvement | 2.5 (1.3-4.7) | **0.004** |
| Large-artery atherosclerosis (LAA) | 1.2 (0.7-2.1) | 0.42 |
| EEG parameters |  |  |
| Generalised slowing | 1.5 (0.9-2.5) | 0.16 |
| Regional slowing | 2.6 (1.4-4.6) | **0.002** |
| All epileptiform activities (IEA, GPDs, LPDs, LRDA, ESz, ESE) | 1.8 (1.1-2.9) | **0.024** |

IEA, sporadic interictal epileptic activity; GPDs, generalised periodic discharges; LPDs, lateralized periodic discharges; LRDA, lateralized rhythmic delta activity; ESz, electrographic seizures; ESE, electrographic status epilepticus

Supplemental Table 5: *Concordance Statistics Summary*

**Concordance Statistics Summary for SeLECT_2.0_ and SeLECT-EEG Models (Derivation Cohorts)**

| **Cohort Type** | **SeLECT_2.0_ (C-Statistic, 95% CI)** | **SeLECT-EEG (C-Statistic, 95% CI)** | **p-value** |
| --- | --- | --- | --- |
| All (Derivation) | 0.69 (0.64-0.74) | 0.73 (0.69-0.78) | **<0.001** |
| No ASyS (Derivation) | 0.71 (0.65-0.75) | 0.75 (0.71-0.80) | **<0.001** |
| ASyS (Derivation) | 0.59 (0.48-0.72) | 0.59 (0.48-0.71) | 0.97 |

**Concordance Statistics Summary for SeLECT_2.0_ and SeLECT-EEG Models (Validation Cohorts)**

| **Cohort Type** | **SeLECT_2.0_ (C-Statistic, 95% CI)** | **SeLECT-EEG (C-Statistic, 95% CI)** | **p-value** |
| --- | --- | --- | --- |
| No ASyS (Validation) | 0.79 (0.59-0.99) | 0.83 (0.66-1.00) | 0.31 |

**SeLECT-EEG Short vs. Continuous Recording in Derivation Cohorts**

| **EEG Type** | **Cohort Type** | **C-Statistic (95% CI)** | **p-value** |
| --- | --- | --- | --- |
| Short EEG | Derivation | 0.72 (0.66-0.77) |  |
| Continuous EEG | Derivation | 0.75 (0.71-0.80) | **<0.001** |

The SeLECT-EEG model consistently outperformed the SeLECT_2.0_ model in both derivation and validation cohorts, particularly among stroke survivors without ASyS, where it showed significantly higher concordance. In derivation cohorts, the SeLECT-EEG model demonstrated a C-Statistic improvement from 0.69 to 0.73 in all stroke survivors and from 0.71 to 0.75 in those without ASyS, both with significant p-values. However, in stroke survivors with ASyS, both models performed similarly, with no significant difference. In validation cohorts, the SeLECT-EEG model also showed better concordance (0.83 vs. 0.79), though the difference was not statistically significant. Additionally, cEEG recordings proved more reliable than short EEGs, showing superior concordance in derivation cohorts.

Supplemental Table 6: *Results of multivariate Cox-regression models of time to first remote symptomatic seizure in stroke survivors with acute-symptomatic seizures (n=108)*

|  | **Cox-regression**  **(full model)** | |
| --- | --- | --- |
| **Variable** | **aHR**  **(95% CI)** | **P value** |
| Stroke severity at admission |  |  |
| NIHSS 4-10 | 0.6 (0.2-1.7) | 0.31 |
| NIHSS ≥11 | 0.8 (0.4-3.0) | 0.82 |
| Stroke location |  |  |
| Middle cerebral artery territory involvement | 1.5 (0.6-3.7) | 0.43 |
| Cortical involvement | 1.0 (0.3-2.6) | 0.91 |
| Large-artery atherosclerosis (LAA) | 1.8 (0.8-4.0) | 0.13 |
| EEG parameters |  |  |
| Generalised slowing | 1.2 (0.5-3.0) | 0.66 |
| Regional slowing | 1.1 (0.5-2.4) | 0.76 |
| All epileptiform activities (IEA, GPDs, LPDs, LRDA, ESz, ESE) | 2.5 (1.1-6.0) | **0.033** |

IEA, sporadic interictal epileptic activity; GPDs, generalised periodic discharges; LPDs, lateralized periodic discharges; LRDA, lateralized rhythmic delta activity; ESz, electrographic seizures; ESE, electrographic status epilepticus

Supplemental Table 7: *Internal Cross-validation Using C-statistics*

| **Omitted cohort** | **Observations**  **(n)** | **SeLECT_2.0_**  **ROC Area (95%-CI)** | **SeLECT-EEG**  **ROC Area (95%-CI)** | **p-value** |
| --- | --- | --- | --- | --- |
| Switzerland (1) | 669 | 0.71 (0.65-0.77) | 0.76 (0.71-0.81) | **<0.001** |
| Switzerland (2) | 615 | 0.67 (0.61-0.73) | 0.71 (0.66-0.77) | **0.002** |
| Portugal | 743 | 0.70 (0.63-0.77) | 0.75 (0.70-0.81) | **<0.001** |
| Belgium | 815 | 0.70 (0.64-0.76) | 0.75 (0.70-0.81) | **<0.001** |
| USA | 646 | 0.75 (0.68-0.81) | 0.77 (0.71-0.83) | **0.05** |
| Mean AUC |  | **0.71** | **0.75** |  |
| Paired t-test |  | t=-7.2029 | **df = 4** | **<0.001** |
| Mean Difference (95% CI) | | -0.042 (-0.058 to -0.026) | | |

The table shows ROC AUC values for the SeLECT2.0 and SeLECT-EEG models across five cohorts using a leave-one-out validation method. Mean AUCs were 0.706 for SeLECT2.0 and 0.748 for SeLECT-EEG, indicating better performance for the latter. A paired t-test (t = -7.2029, p = 0.001969) confirmed a significant improvement with the SeLECT-EEG model. The 95% confidence interval for the mean difference in AUCs was -0.058 to -0.026, highlighting the superior calibration and discrimination of the SeLECT-EEG model.

Supplemental Table 8: *Patient and clinical characteristics, stratified by groups with regularly in clinical setting conducted EEG (Switzerland (1) and (2), Portugal) vs. selected stroke survivors for cEEG (group 2: Belgium and USA) in stroke survivors without acute symptomatic seizures (n=872, EEG variables for short EEG 20-60min)*

| **Variable** | **Group 1**  **(Switzerland (1) and (2), Portugal)** | **Group 2**  **(Belgium, USA)** | **OR or MD**  **(95% CI)** | **P-value** |
| --- | --- | --- | --- | --- |
| Total | 589 | 283 |  |  |
| Age *(years)* | 71 (57-84) | 64 (50-78) | -7 (-8.9 - -5.1) | **< 0.001** |
| Sex |  |  |  |  |
| Male | 320 (54%) | 144 (51%) | 0.9 (0.7 - 1.2) | 0.35 |
| Female | 269 (46%) | 139 (49%) | 1.1 (0.9 - 1.5) | 0.35 |
| Time of follow-up (in months) | 22 (12-47) | 41 (22-66) | 19.0 (15.5 - 22.5) | **< 0.001** |
| NIHSS at admission |  |  |  |  |
| ≤3 | 218 (37%) | 88 (21%) | 0.8 (0.6 - 1.0) | 0.10 |
| 4-10 | 221 (37%) | 78 (20%) | 0.6 (0.5 - 0.9) | **0.004** |
| ≥11 | 150 (26%) | 117 (58%) | 2.1 (1.5 - 2.8) | **<0.001** |
| Stroke location |  |  |  |  |
| Middle cerebral artery territory involvement | 461 (78%) | 214 (76%) | 0.9 (0.6 - 1.2) | 0.39 |
| Cortical involvement | 351 (60%) | 196 (69%) | 1.5 (1.1 - 2.1) | **0.006** |
| Stroke cause |  |  |  |  |
| Large-artery atherosclerosis | 91 (22%) | 58 (21%) | 1.4 (1.0 - 2.0) | 0.07 |
| All other | 498 (78%) | 225 (79%) | 0.7 (0.5 - 1.0) | 0.07 |
| Clinical seizures |  |  |  |  |
| Late Seizures | 36/589 (6.1%) | 41/283 (14%) | 2.6 (1.6 - 4.2) |  |
| EEG parameters |  |  |  |  |
| Normal | 165/589 (28%) | 35/283 (12%) | 0.4 (0.2 - 0.5) | **<0.001** |
| Generalised slowing | 278/589 (14%) | 178/283 (63%) | 1.9 (1.4 - 2.5) | **< 0.001** |
| Generalised Rhythmic Slowing | - | 35/283 (12.4%) | - |  |
| Regional slowing | 285/589 (48%) | 115/283 (41%) | 0.7 (0.5 - 1.0) | **0.04** |
| Regional rhythmic slowing (LRDA) | - | 13/283 (4.6%) | - |  |
| Generalised periodic discharges | - | 6/283 (1.1%) | - |  |
| Sporadic interictal epileptic discharges/activity (IEA) | 24/589 (4.1%) | 9/226 (4%) | 0.8 (0.4 - 1.7) | 0.58 |
| Lateralized periodic discharges (LPDs) | 31/589 (5.3%) | 6/283 (2.1%) | 0.4 (0.2 - 0.9) | **0.03** |
| IEA or LPDs | 31/589 (7.5%) | 15/283 (5.3%) | 1.0 (0.5 - 1.9) | 1.0 |
| IEA or LPDs or LRDA | 44/589 (7.5%) | 27/283 (9.5%) | 1.3 (0.8 - 2.2) | 0.29 |
| Electrographic seizures (ESz) | 0/589 (0%) | 2/283 (0.7%) | - |  |
| Electrographic status epilepticus (ESE) | 1/589 (0.2%) | 1/283 (0.4%) | 2.1 (0.1 - 33.5) | 0.54 |
| All epileptiform activity (IEA, GPDs, LPDs, LRDA, ESz, ESE) | 44/589 (7.5%) | 33/283 (12%) | 1.6 (1.0-2.6) | 0.06 |

This table displays the odds ratios (OR) and mean differences (MD) comparing Group 2 (Belgium, USA) to Group 1 (Switzerland, Portugal). The ORs are calculated from the perspective of Group 2, with significant findings highlighted. Mean differences are utilized for continuous variables such as age and time of follow-up. Notably, Group 2 had increased odds for high NIHSS scores (OR 2.1, 95% CI: 1.5-2.8, p<0.001) and generalised slowing (OR 1.9, 95% CI: 1.4-2.5, p<0.001), but lower odds for regional slowing (OR 0.7, 95% CI: 0.5-1.0, p=0.04) and lateralized periodic discharges (LPD) (OR 0.8, 95% CI: 0.4-1.7, p=0.03). This suggests that the high NIHSS scores and resulting generalised slowing in Group 2 may have complicated the detection of other electrographic abnormalities. Additionally, Group 2 had significantly higher odds for cortical involvement (OR 1.5, 95% CI: 1.1-2.1, p=0.006) and late seizures (OR 2.6, 95% CI: 1.6-4.2), while the mean age was significantly lower (MD: -7 years, 95% CI: -8.9 to -5.1, p<0.001) and the time of follow-up significantly longer (MD: 19 months, 95% CI: 15.5-22.5, p<0.001).

Supplemental Table 9: *Mediation Analysis of EEG Variables on Late Seizure Outcomes (n=980)*

| **Ind**epe**ndent Variable** | **Mediator** | **ACME** | | **ADE** | | **Total Effect** | | **Prop. Mediated**  **in %** |
| --- | --- | --- | --- | --- | --- | --- | --- | --- |
|  |  | **Value (95%-CI)** | **p-value** | **Value (95%-CI)** | **p-value** | **Value (95%-CI)** | **p-value** |  |
| Regional slowing | Generalised slowing | 0.014  (0.006-0.023) | **<0.001** | 0.086  (0.050-0.125) | **<0.001** | 0.100  (0.063-0.142) | **<0.001** | **14** |
| Regional slowing | Epileptiform Activity | 0.030  (0.017-0.045) | **<0.001** | 0.070  (0.034-0.110) | **0.002** | 0.100  (0.064-0.141) | **<0.001** | **30** |
| Generalised slowing | Regional slowing | 0.010  (0.003-0.18) | **<0.001** | 0.127  (0.081-0.179) | **<0.001** | 0.137  (0.092-0.187) | **<0.001** | **7** |
| Generalised slowing | Epileptiform Activity | 0.042  (0.024-0.065) | **<0.001** | 0.095  (0.054-0.139) | **<0.001** | 0.137  (0.093-0.183) | **<0.001** | **30** |
| Epileptiform Activity | Regional slowing | 0.021  (0.009-0.035) | **<0.001** | 0.198  (0.123-0.280) | **<0.001** | 0.219  (0.144-0.299) | **<0.001** | **9** |
| Epileptiform Activity | Generalised slowing | 0.040  (0.022-0.061) | **<0.001** | 0.178  (0.107-0.252) | **<0.001** | 0.219  (0.149-0.294) | **<0.001** | **19** |

IEA, sporadic interictal epileptic activity; GPDs, generalised periodic discharges; LPDs, lateralized periodic discharges; LRDA, lateralized rhythmic delta activity; ESz, electrographic seizures; ESE, electrographic status epilepticus

The mediation analysis reveals that epileptiform activity is a significant mediator in the relationship between EEG variables and late seizure outcomes. Regional slowing, when mediated by epileptiform activity, has an ACME of 0.030 (95%-CI: 0.017-0.045, p < 0.001), contributing to 30% of the total effect, while generalised slowing mediated by epileptiform activity shows an ACME of 0.042 (95%-CI: 0.024-0.065, p < 0.001), also accounting for 30% of the total effect. In contrast, the mediation effects of generalised slowing by regional slowing and vice versa were lower, with ACME values of 0.010 (95%-CI: 0.003-0.018, p < 0.001) and 0.014 (95%-CI: 0.006-0.023, p < 0.001), contributing 7% and 14% of the total effect, respectively. These findings underscore the critical role of epileptiform activity in influencing late seizure outcomes.

### Supplemental Figure 1: Flowchart of Post-Stroke Survivors with Available Short and/or cEEG and Subsequent Cohort Derivation and Validation

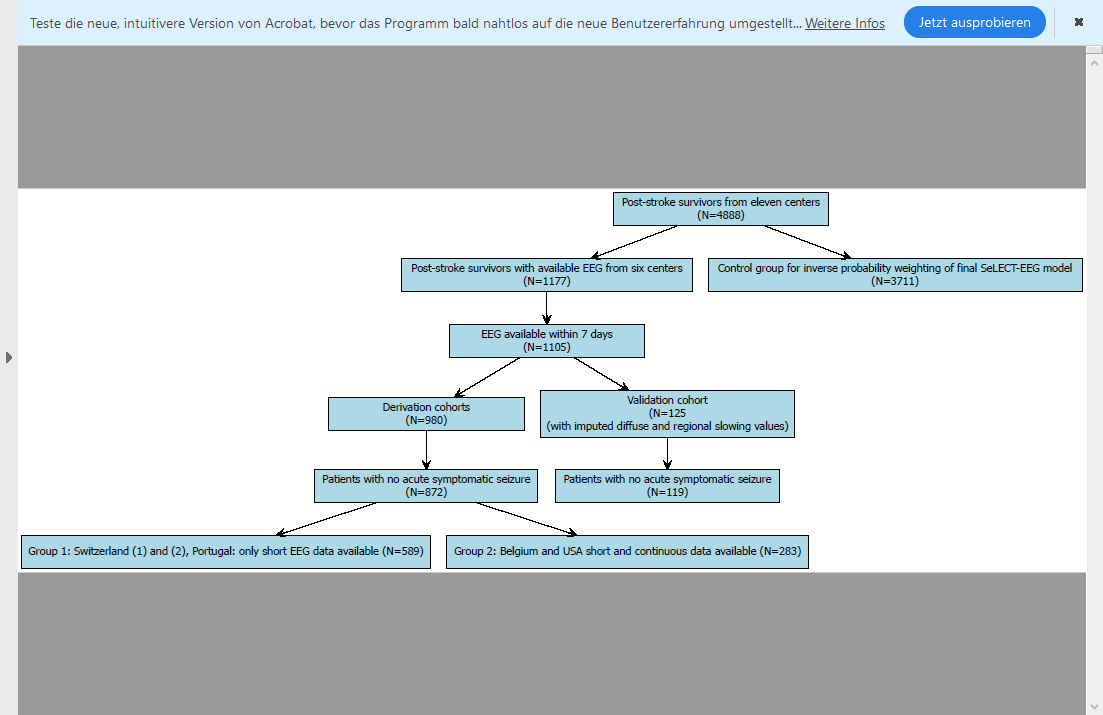

Supplemental Figure 2: *Calibration plots for old SeLECT_2.0_ and new SeLECT-EEG model in derivation and validation cohorts*

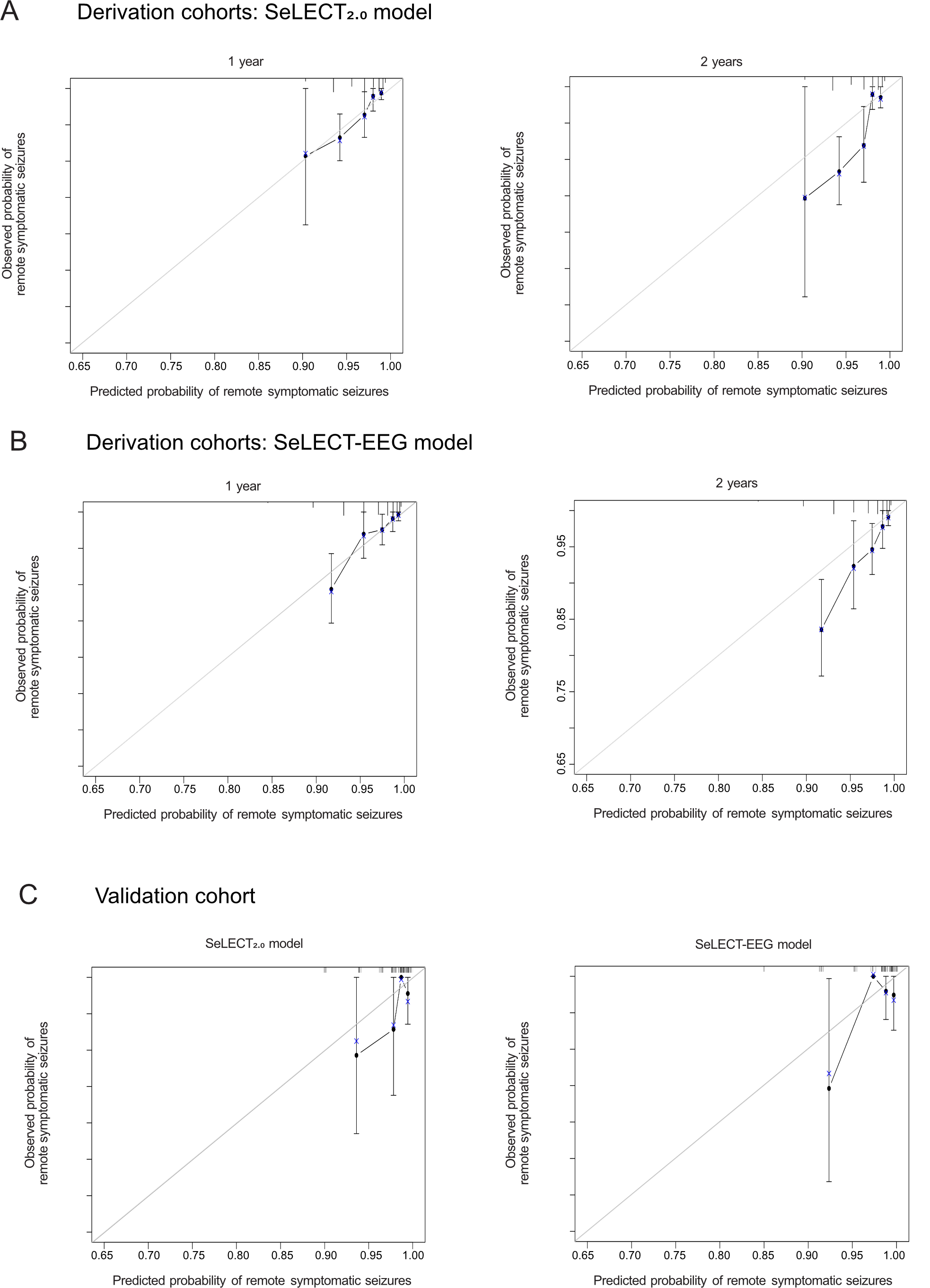
 Calibration plots for the SeLECT2.0 model (Panel A) and the SeLECT-EEG model (Panel B) in the derivation cohort at 1-year (left) and 2-years (right) post-stroke onset, using a Kaplan-Meier calibration method. Panel C presents the calibration plots for the SeLECT_2.0_ model (left) and the SeLECT-EEG model (right). Vertical lines represent 95% confidence intervals. The calibration plots indicate high agreement between predicted and observed outcomes for both models in both the derivation and validation cohorts. Brier scores for the derivation cohort were 0.0783 for SeLECT_2.0_ and 0.0770 for SeLECT-EEG, while for the validation cohort they were 0.0253 for SeLECT_2.0_ and 0.0241 for SeLECT-EEG. The t-test results showed a non-significant trend towards improved calibration with the SeLECT-EEG model. Despite strong overall calibration, both models show challenges in accurately predicting lower risk groups due to low event rates.

Supplemental Figure 3: *EEG findings for short (20-60min) and continuous (>12h) EEG recordings*

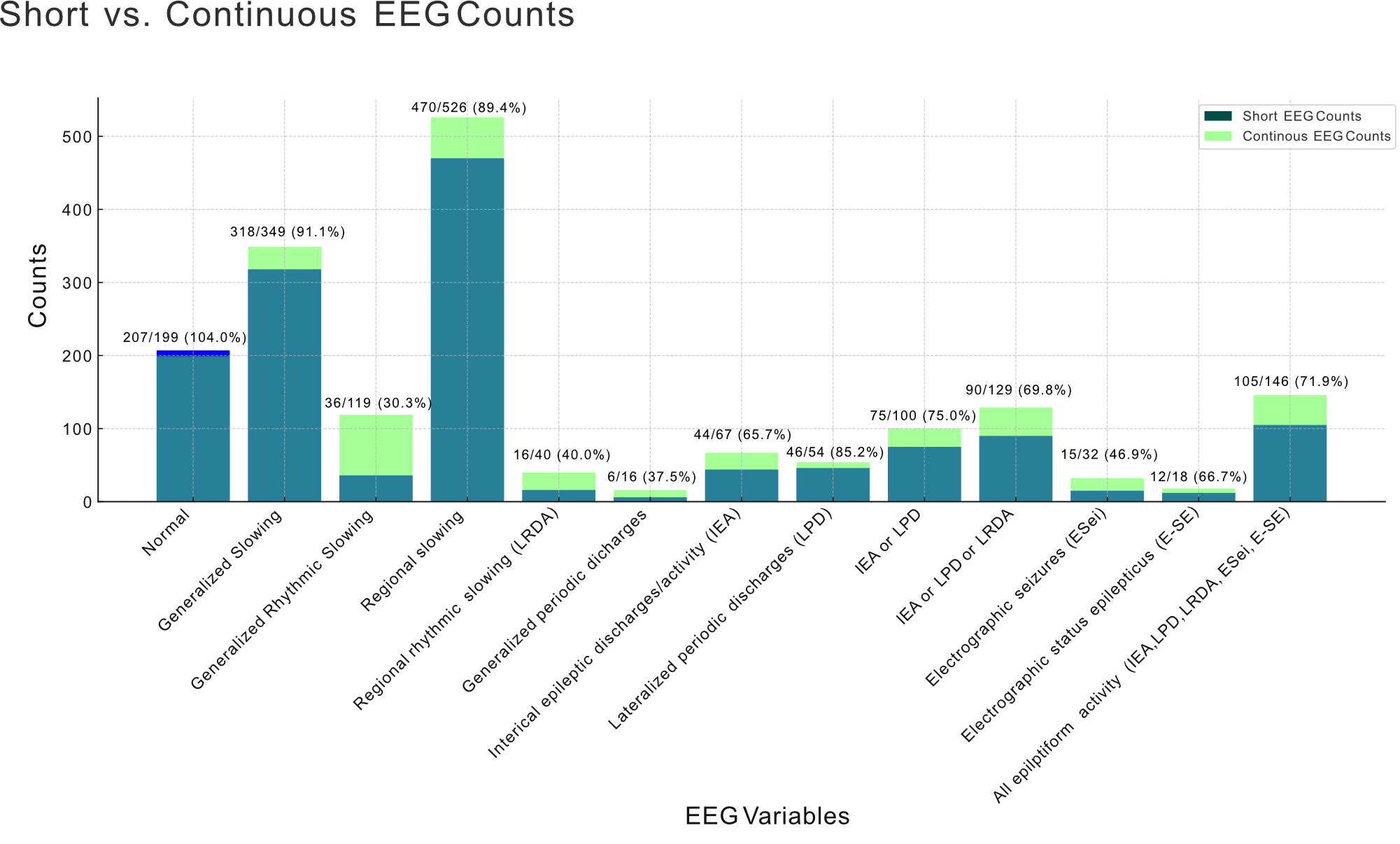

Detection rates of various electrographic findings in short EEGs (20-60 minutes) versus cEEGs (at least >12 hours). The counts of each finding type are displayed, with the percentage of findings in short EEGs relative to cEEGs annotated above each bar. Overall, some findings, such as normal patterns as well as generalised and regional slowing, had high detection rates in short EEGs, comparable to cEEGs. However, other findings, like epileptiform activity and rhythmic and periodic patterns, showed significantly lower detection rates in short EEGs.

Supplemental Figure 4: *Impact of different SeLECT-EEG score values and seizure-free intervals on the chance of an occurrence of a seizure in the next year*

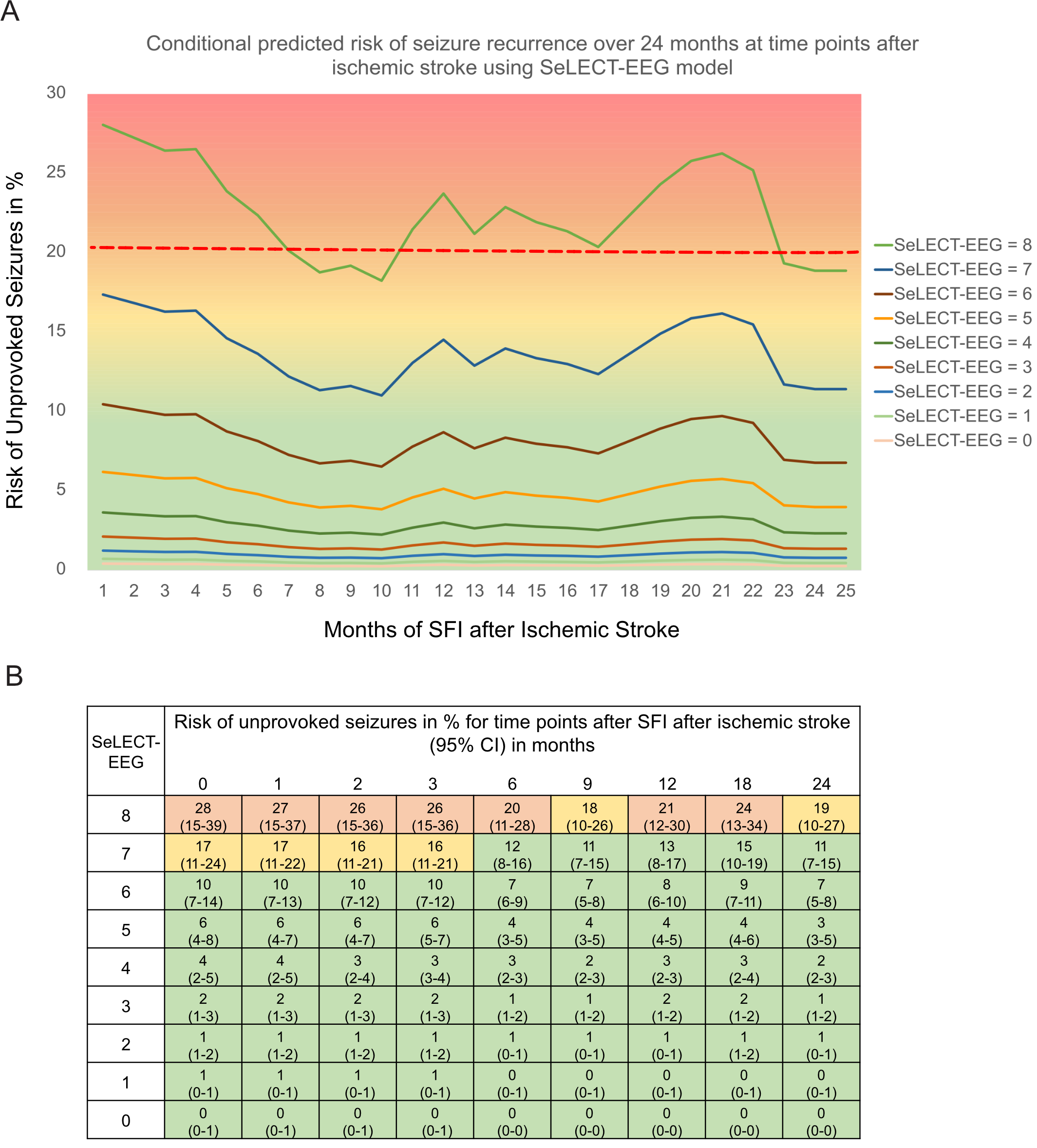
 **Panel A** displays the impact of different seizure-free intervals (SFI) on the chance of an occurrence of a seizure in the next year (COSY) following ischemic stroke. The lines represent different SeLECT-ASyS scores. **Panel B** shows the numerical estimates of COSY stratified by different SFIs and SeLECT-EEG values including the 95% confidence intervals. Colors suggested by different approaches by Bonnett et al.^6^ and Marson^7^ (acceptable range of risk for private driving for COSY of 20-40% suggested by Schmedding^8^): risk estimates ≥ 20% (and lower CI ≥ 20%) in red ("permissive approach"); risk estimates < 20% (and higher CI < 20%) in green ("conservative approach") in orange and yellow: orange when risk estimate ≥ 20% but lower CI < 20% (= "liberal approach"); yellow when risk estimate < 20% but upper CI > 20% (=" intermediate approach").

Supplemental Figure 5: *Predicted Risk of Post-Stroke Epilepsy according to a new prognostic model in stroke survivors without acute symptomatic seizures including electrographic findings*

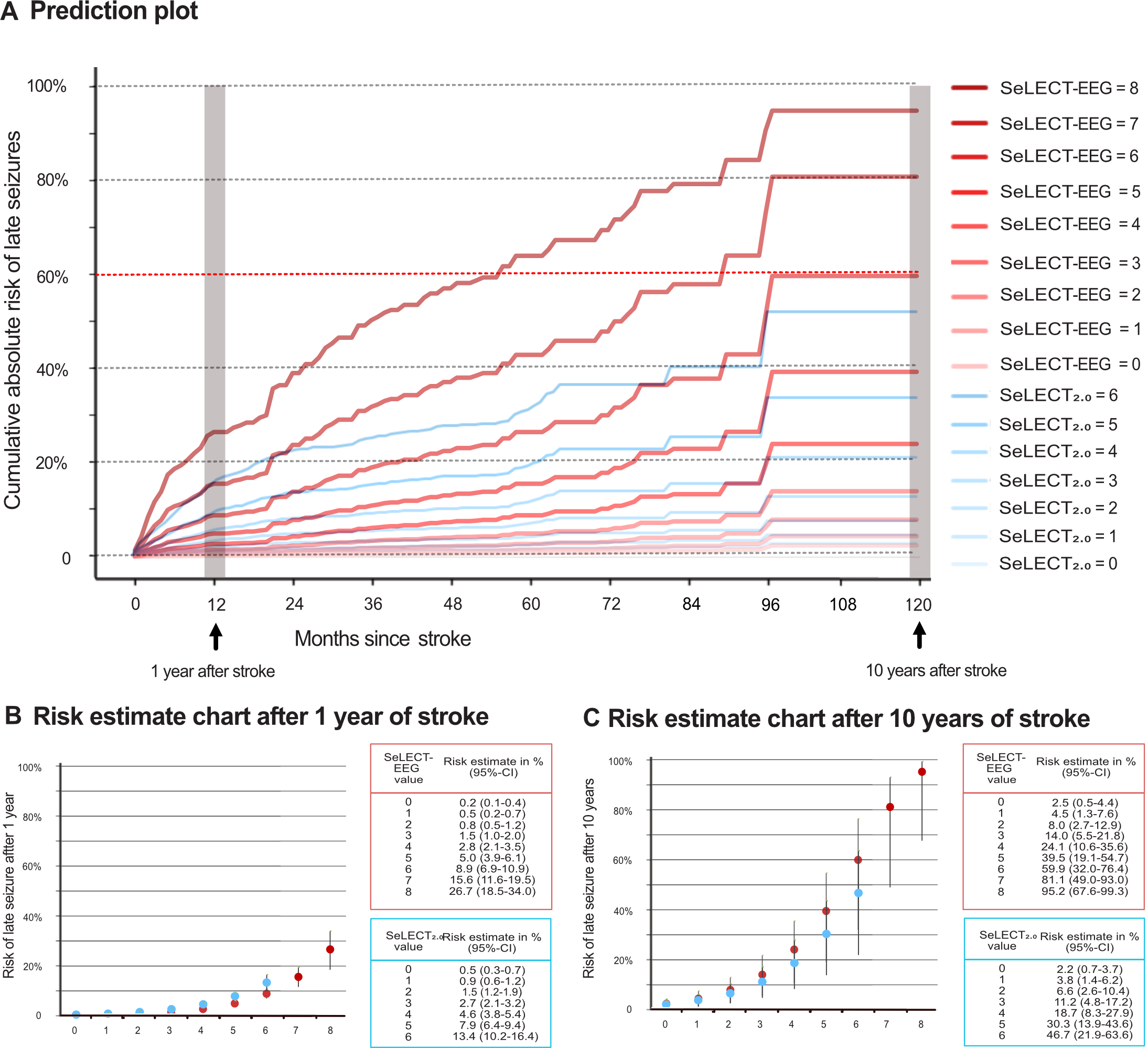

**Panel A** illustrates the predicted risk of unprovoked remote symptomatic seizures occurring 0-120 months post-stroke. The red curves represent SeLECT-EEG values for stroke survivors including electrographic findings, while the blue curves represent the prior SeLECT_2.0_ model for stroke survivors without these abnormalities. Each curve corresponds to SeLECT-EEG values ranging from 0-8 (in red) and SeLECT_2.0_ values ranging from 0-6 (in blue).

**Panels B and C** present risk estimate charts for late seizures at 1 year and 10 years post-stroke, respectively, based on the SeLECT-ASyS and SeLECT_2.0_ scores. Vertical lines denote the 95% confidence intervals.

### Author List and Affiliations of the SeLECT Consortium

| **Author** | **Affiliation** |
| --- | --- |
| Dominik Zieglgänsberger, MD | Department of Neurology, Kantonsspital St. Gallen, St Gallen, Switzerland |
| Giulio Bicciato, MD | Department of Neurology, Clinical Neuroscience Center, University Hospital and University of Zurich, Zurich, Switzerland |
| Laura Abraira, MD PhD | Epilepsy Unit, Department of Neurology, Vall d’Hebron Hospital Universitari, Barcelona; Universitat Autonoma de Barcelona, Bellaterra, Spain |
| Estevo Santamarina, MD | Epilepsy Unit, Department of Neurology, Vall d’Hebron Hospital Universitari, Barcelona; Universitat Autonoma de Barcelona, Bellaterra, Spain |
| José Álvarez-Sabín, PhD | Epilepsy Unit, Department of Neurology, Vall d’Hebron Hospital Universitari, Barcelona; Universitat Autonoma de Barcelona, Bellaterra, Spain |
| Carolina Ferreira-Atuesta, MD MSc | Department of Clinical & Experimental Epilepsy, UCL Queen Square Institute of Neurology, London WC1N 3BG & Chalfont Centre for Epilepsy, Chalfont St Peter SL9 0RJ, United Kingdom; Department of Neurology, Icahn School of Medicine at Mount Sinai, New York, United States |
| Mira Katan, MD MSc | Department of Neurology, Clinical Neuroscience Center, University Hospital and University of Zurich, Zurich, Switzerland; Department of Neurology, University Hospital and University of Basel, Basel, Switzerland |
| Natalie Scherrer, MD | Department of Neurology, Clinical Neuroscience Center, University Hospital and University of Zurich, Zurich, Switzerland |
| Robert Terziev, MD | Department of Neurology, Clinical Neuroscience Center, University Hospital and University of Zurich, Zurich, Switzerland |
| Nico Döhler, MD | Department of Neurology, Kantonsspital St. Gallen, St Gallen, Switzerland; Specialist Clinic for Neurorehabilitation, Kliniken Beelitz, Beelitz-Heilstätten, Germany |
| Barbara Erdélyi-Canavese, MD | Department of Neurology, Kantonsspital St. Gallen, St Gallen, Switzerland |
| Ansgar Felbecker, MD | Department of Neurology, Kantonsspital St. Gallen, St Gallen, Switzerland |
| Philip Siebel, MD | Department of Neurology, Kantonsspital St. Gallen, St Gallen, Switzerland |
| Michael Winklehner, MD | Johannes Kepler University Linz, Kepler University Hospital, Department of Neurology, Altenberger Straße 69, 4040 Linz and Wagner-Jauregg Weg 15, 4020 Linz, Austria |
| Tim J von Oertzen, MD FRCP | Johannes Kepler University Linz, Kepler University Hospital, Department of Neurology, Altenberger Straße 69, 4040 Linz and Wagner-Jauregg Weg 15, 4020 Linz, Austria |
| Judith N. Wagner, MD | Johannes Kepler University Linz, Kepler University Hospital, Department of Neurology, Altenberger Straße 69, 4040 Linz and Wagner-Jauregg Weg 15, 4020 Linz, Austria; Department of Neurology, Evangelisches Klinikum Gelsenkirchen, Academic Hospital University Essen-Duisburg, Gelsenkirchen, Germany |
| Gian Luigi Gigli, MD | Department of Medicine, University of Udine and Clinical Neurology, Udine University Hospital, Udine, Italy |
| Annacarmen Nilo, MD | Department of Medicine, University of Udine and Clinical Neurology, Udine University Hospital, Udine, Italy |
| Francesco Janes, MD PhD | Department of Medicine, University of Udine and Clinical Neurology, Udine University Hospital, Udine, Italy |
| Giovanni Merlino, MD PhD | Department of Medicine, University of Udine and Clinical Neurology, Udine University Hospital, Udine, Italy |
| Mariarosaria Valente, MD | Department of Medicine, University of Udine and Clinical Neurology, Udine University Hospital, Udine, Italy |
| María Paula Zafra-Sierra, MD | Department of Neurology, Fundación Santa Fe de Bogotá, Universidad de Los Andes, Universidad del Bosque, Bogotá, Colombia |
| Luis Carlos Mayor-Romero, MD | Department of Neurology, Fundación Santa Fe de Bogotá, Universidad de Los Andes, Universidad del Bosque, Bogotá, Colombia |
| Julian Conrad, MD | Department of Neurology, University of Muenster, Muenster, Germany; Division for neurodegenerative diseases, Department of Neurology, Universitaetsmedizin Mannheim, University of Heidelberg |
| Stefan Evers, MD PhD | Department of Neurology, University of Muenster, Muenster, Germany; Department of Neurology, Krankenhaus Lindenbrunn, Coppenbrügge, Germany |
| Piergiorgio Lochner, MD | Department of Neurology, Saarland University Medical Center, Homburg, Germany |
| Frauke Roell, MD | Department of Neurology, Saarland University Medical Center, Homburg, Germany |
| Francesco Brigo, MD | Department of Neurology, Hospital of Merano (SABES-ASDAA), Merano-Meran, Italy |
| Mark R Keezer, MDCM PhD | Department of Clinical & Experimental Epilepsy, UCL Queen Square Institute of Neurology, London WC1N 3BG & Chalfont Centre for Epilepsy, Chalfont St Peter SL9 0RJ, United Kingdom; Centre Hospitalier de l’Université de Montréal, Montreal, QC, Canada |
| John S Duncan, FRCP FMedSci | Department of Clinical & Experimental Epilepsy, UCL Queen Square Institute of Neurology, London WC1N 3BG & Chalfont Centre for Epilepsy, Chalfont St Peter SL9 0RJ, United Kingdom |
| Josemir W Sander, FRCP FMedSci | Department of Clinical & Experimental Epilepsy, UCL Queen Square Institute of Neurology, London WC1N 3BG & Chalfont Centre for Epilepsy, Chalfont St Peter SL9 0RJ, United Kingdom; Stichting Epilepsie Instellingen Nederland –(SEIN), Heemstede 2103 SW, The Netherlands |
| Barbara Tettenborn, MD | Department of Neurology, Kantonsspital St. Gallen, St Gallen, Switzerland |
| Matthias J Koepp, MD PhD | Department of Clinical & Experimental Epilepsy, UCL Queen Square Institute of Neurology, London WC1N 3BG & Chalfont Centre for Epilepsy, Chalfont St Peter SL9 0RJ, United Kingdom |
